# Relative Ratios of Human Seasonal Coronavirus Antibodies Predict the Efficiency of Cross-Neutralization of SARS-CoV-2 Spike Binding to ACE2

**DOI:** 10.1101/2021.07.16.21260079

**Authors:** Yannick Galipeau, Vinayakumar Siragam, Geneviève Laroche, Erika Marion, Matthew Greig, Michaeline McGuinty, Ronald A Booth, Yves Durocher, Miroslava Cuperlovic-Culf, Steffany A.L. Bennett, Angela M. Crawley, Patrick M. Giguère, Curtis Cooper, Marc-André Langlois

## Abstract

**Background:** Antibodies raised against human seasonal coronaviruses (sCoVs), which are responsible for the common cold, are known to cross-react with SARS-CoV-2 antigens. This prompts questions about their protective role against SARS-CoV-2 infections and COVID-19 severity. However, the relationship between sCoV exposure and SARS-CoV-2 correlates of protection are not clearly identified.

**Methods:** We performed a cross-sectional analysis of cross-reactivity and cross-neutralization to SARS-CoV-2 antigens (S-RBD, S-trimer, N) using pre-pandemic serum from four different groups: pediatrics and adolescents, persons 21 to 70 years of age, older than 70 years of age, and persons living with HCV or HIV.

**Findings:** Antibody cross-reactivity to SARS-CoV-2 antigens varied between 1.6% and 15.3% depending on the cohort and the isotype-antigen pair analyzed. We also show a range of neutralizing activity (0-45%) in serum that interferes with SARS-CoV-2 spike attachment to ACE2. While the abundance of sCoV antibodies did not directly correlate with neutralization, we show that neutralizing activity is rather dependent on relative ratios of IgGs in sera directed to all four sCoV spike proteins. More specifically, we identified antibodies to NL63 and OC43 as being the most important predictors of neutralization.

**Interpretation:** Our data support that exposure to sCoVs triggers antibody responses that influence the efficiency of SARS-CoV-2 spike binding to ACE2, and may also impact COVID-19 disease severity through other latent variables.

**Research in Context:** *Evidence before this study:* There is a growing body of evidence showing that within the population there are varying levels of pre-existing immunity to SARS-CoV-2 infection and possibly COVID-19 disease severity. This immunity is believed to be attributable to prior infection by four prevalent seasonal coronaviruses (sCoVs) responsible for the common cold. Pre-existing immunity can be assessed in part by antibodies directed to sCoVs that also cross-react to SARS-CoV-2 antigens. The SARS-CoV-2 spike and, more specifically, the receptor binding domain are the primary targets for neutralizing antibodies. It is unclear if cross-reactive antibodies to SARS-CoV-2 are neutralizing and are also responsible for the broad spectrum of COVID-19 disease severity, from asymptomatic to critical, observed in the infected population.

*Added-value of this study:* Here we carried out a detailed analysis of sCoV prevalence in samples acquired before the pandemic from individuals of various age groups and in people living with HIV and HCV. We then analyzed the frequency of all the different types of antibodies that cross-react to three SARS-CoV-2 antigens. We found a high level of people with cross-reactive antibodies, surprisingly we also detected that some people have antibodies that block the SARS-CoV-2 spike from binding to its human receptor, ACE2. By using machine learning, we were able to accurate predict which individuals can neutralize SARS-CoV-2 spike-ACE2 interactions based on their relative ratios of antibodies against the four sCoVs.

*Implications of all the available evidence:* We demonstrate that it not absolute levels of sCoVs antibodies that are predictive of neutralization but the relative ratios to all four sCoVs, with NL63 being the most weighted for this prediction. Machine learning also highlighted the existence of latent variables that contribute to the neutralization and that may be related to the type of cellular immune response triggered by the infection to certain sCoVs. This study is one of the first to identify a functional relationship between prior-exposure to sCoV and the establishment of a certain degree of immunity to SARS-CoV-2 by way of a cross-reactive antibody response.

**Graphical Abstract:** 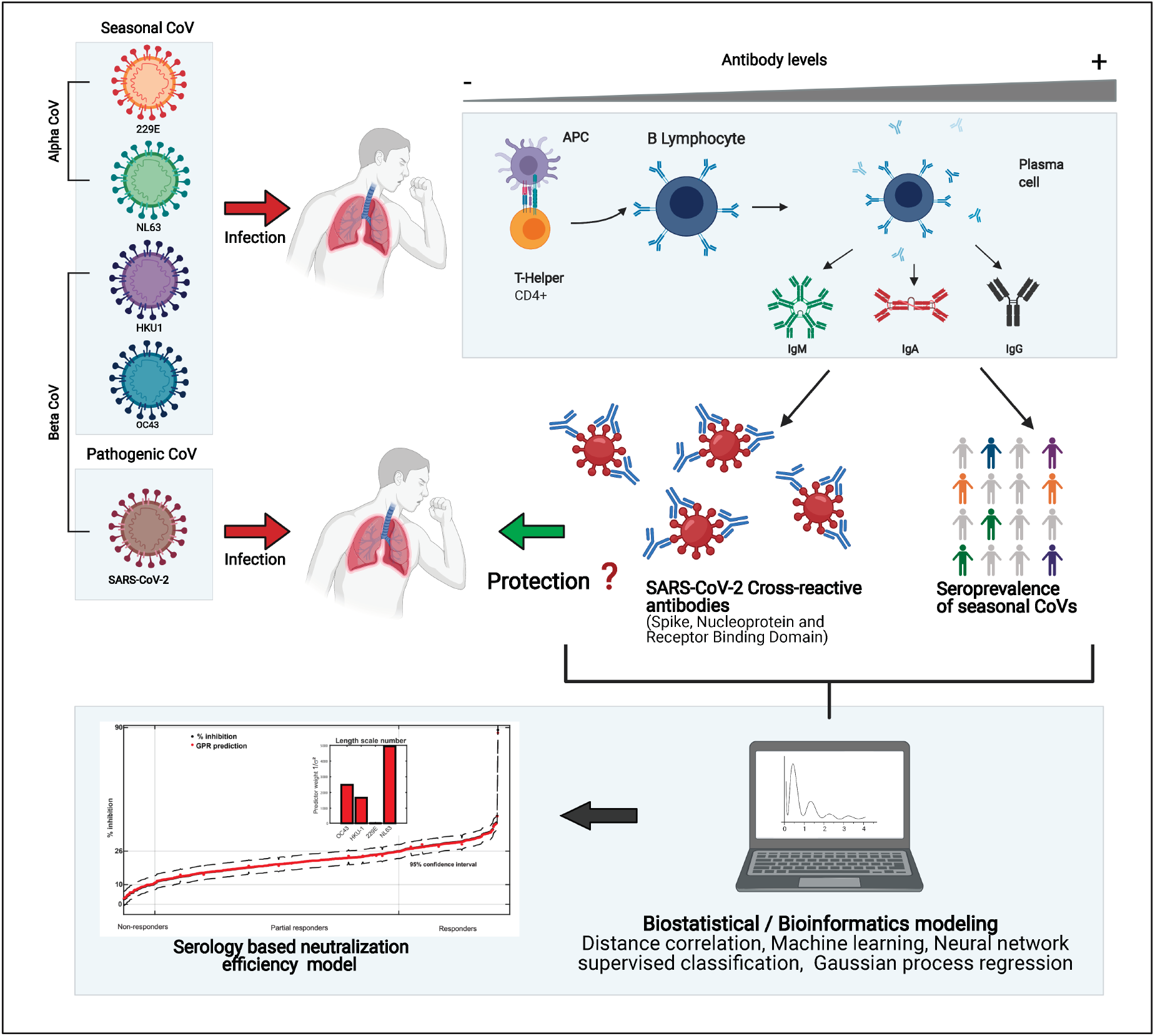

## Introduction

Four endemic human sCoVs (229E, OC43, NL63, and HKU1) are highly prevalent worldwide and cause common and recurrent respiratory infections (1–3). A retrospective prevalence study of acute respiratory infections in France found that OC43 is the most prevalent sCoV followed by NL63, HKU1, and finally 229E (4). While nearly every adult has been exposed and demonstrates humoral responses to one or several of these sCoV (5–7), immunity to each specific sCoV wanes over time. A 35-year longitudinal study revealed that reinfections by a same sCoV were regularly observed at 6 and 9 months post-infection, but most were most frequently observed after 12 months (3). These findings highlight that sterilizing immunity to these viruses is short-lived and that the relative ratios of antibodies against any one of the four sCoVs is highly variable and dependent on the most recent exposure. This study also indicates that exposure to one sCoV does not offer complete protection against infection by the other viruses, but it is unclear how prior immunity affects disease severity.

Seasonal CoVs generally share an overall low degree of sequence similarities with SARS-CoV-2 proteins and only the alphacoronavirus NL63 also utilizes ACE2 as an entry receptor (8, 9). Nevertheless, conserved epitopes on the spike S2 domain are believed to be responsible for antibody and T-cell cross reactivity between sCoVs and other human betacoronaviruses such as SARS-CoV-1, SARS-CoV-2 and MERS (10–14). Furthermore, the nucleoprotein (N) is one of the most conserved antigens across all human CoVs (15). In fact, antibodies against sCoV antigens impede the specificity of serological tests against SARS-CoV-2 when full length S-trimer and N antigens are used in the assay (reviewed here: (16)). While neutralizing activity directed against the SARS-CoV-2 S protein, more specifically the receptor binding domain (S-RBD), have been extensively described, non-RBD neutralizing epitopes have also been identified on both the S1 and S2 subunits (17–20). Furthermore, antibodies can exhibit non-neutralizing effector functions such as ADCC and complement-fixing that restrict SARS-CoV-2 infection by binding to viral protein epitopes expressed on the surface of infected cells, including S and N antigens (21–24).

While antibody and T-cell responses of SARS-CoV-2 convalescent individuals have been well characterized (24–26), the influence of pre-existing immunity, both humoral and cellular, from exposure to sCoVs is not yet clearly established. While several recent studies have investigated the impact of prior immunity to sCoVs on SARS-CoV-2 infection, neutralization, and disease severity (1, 6, 10, 12, 27–31), there is some discordance in the findings. Some studies have shown weak or no cross-protective antibodies in pre-pandemic blood of donors *in vitro* (1, 6, 31), while several other studies have demonstrated various lines of evidence for antibody and T cell cross-neutralization, and even protection against COVID-19 disease severity (10, 14, 25, 29, 32–34).

Here, we assessed the seroprevalence of sCoVs, along with the neutralization activity and cross-reactivity to the S-trimer, S-RBD and N of SARS-CoV-2, in pre-COVID-19 pandemic sera across four study groups. Our results provide insight into population subgroup variations in the seroprevalence of the four sCoVs, antibody isotype-specific cross-reactivity to SARS-CoV-2 spike (S-trimer), nucleocapsid (N), the receptor binding domain (S-RBD). We also assessed the neutralizing potential of antibodies in pre-pandemic serum that interfere with S binding to the ACE2 receptor. While neutralization of S-trimer binding to ACE2 was detected to various degrees, this activity remained low compared to the neutralizing potential resulting from SARS-CoV-2 infection. However, using several machine learning approaches, we were able to identify a functional dependence on prior sCoV exposure that includes both directly measured and latent variables. These enabled us to model neutralization via Gaussian process regression (GPR) of sCoV seroprevalence. In agreement to previous studies, we do not find a direct correlation between sCoV and neutralization (1, 32, 35). However, we do identify a strong predictive correlation between neutralization of S binding and relative ratios of the different sCoV antibodies. These data support the idea that latent variables associated with sCoV exposure have a predictive and protective role against SARS-CoV-2 infection and possibly disease severity.

## Materials and Methods

### Ethics statement

Use of human samples for this study was approved by the University of Ottawa Ethics Review Board: Certificates H-04-20-5727, H-04-21-6643 and H-07-20-6009. Informed consent was provided by all participants.

### Pre-pandemic cohort

The current study collected pre-pandemic serum or plasma samples separated into four cohorts for a total of 580 enrolled patients. The cohorts selected for inclusion in this study are described in (Table 1), any serum or plasma samples collected prior to SARS-CoV-2 spread in Ottawa with corresponding demographic information was included. Specimens with insufficient volume (<100µL) were excluded. Serum and plasma samples were sourced from diverse source including the Eastern Ontario Regional Laboratory Association (EORLA), the Ottawa Hospital (TOH), and the Icahn School of Medicine at Mount Sinai. Specimens were collected between April 1 2015 and March 4 2020. All the pre-pandemic patient samples were obtained prior to the COVID-19 pandemic in Ottawa. Pediatric samples were acquired from the BC Children’s Hospital Biobank (BCCHB) in Vancouver, BC, Canada (REB#: H-07-20-6009). Due to potential impact of immunosuppression and chronic viral infection on seroprevalence of sCoV antibodies and cross-reactivity, sera from individuals living with HIV and HCV was obtained from The Ottawa Hospital (TOH) and Ottawa Hospital Research Institute (OHRI). The patient’s demographic information and clinical outcomes were obtained from hospital records.

**Table 1.** Demographic information of each pre-pandemic cohort. Serum and plasma samples collected pre-2019 were organized in four distinct cohorts. Basic demographic information was collected and compiled from patient files.

| **Cohorts** | **# of Subjects** | **Age range**<br>**(yrs)** | **Female %**<br>**(Mean Age ± SD)** | **Male %**<br>**(Mean Age ± SD)** |
| --- | --- | --- | --- | --- |
| **< 21 yrs** | 193 | 1-20 yrs | 40%<br>(9 ± 6 yrs) | 60%<br>(8 ± 5 yrs) |
| **21-70 yrs** | 273 | 21-70 yrs | 55%<br>(51 ± 14 yrs) | 45%<br>(53 ± 12 yrs) |
| **> 70 yrs** | 64 | 71-97 yrs | 56%<br>(80 ± 7 yrs) | 44%<br>(79 ± 7 yrs) |
| **HCV & HIV** | 50 | 18-80 yrs | 41%<br>(51 ± 18 yrs) | 59%<br>(57 ± 13 yrs) |

Abbreviations: SD, standard deviation

### Pandemic samples

As a reference group for the neutralization and seasonal experiments, serum samples from individuals post-2019 were used. Samples were acquired within several different research studies and separated by their provenance. The control group for the neutralization (Fig. 5) were SARS-CoV-2 positive and hospitalized individuals (mild and severe) and were further separated by day post positive PCR test. The individuals for reference group in Figure 1 were SARS-CoV-2 positive convalescent individuals. The antibody levels of this group were monitored using the same protocol and platform used to assess the seroprevalence of sCoVs but probed using SARS-CoV-2 S-trimer. The negative reference group was comprised of individuals with no history of SARS-CoV-2 infection who are currently enrolled in an ongoing SARS-CoV-2 surveillance study. Basic demographic information was collected and combined in Table S1. Any serum samples acquired within the mentioned studies with corresponding informed consent and demographic information were eligible. Vaccinated individuals and samples with no clear identification or low volume were excluded.

**Figure 1.**
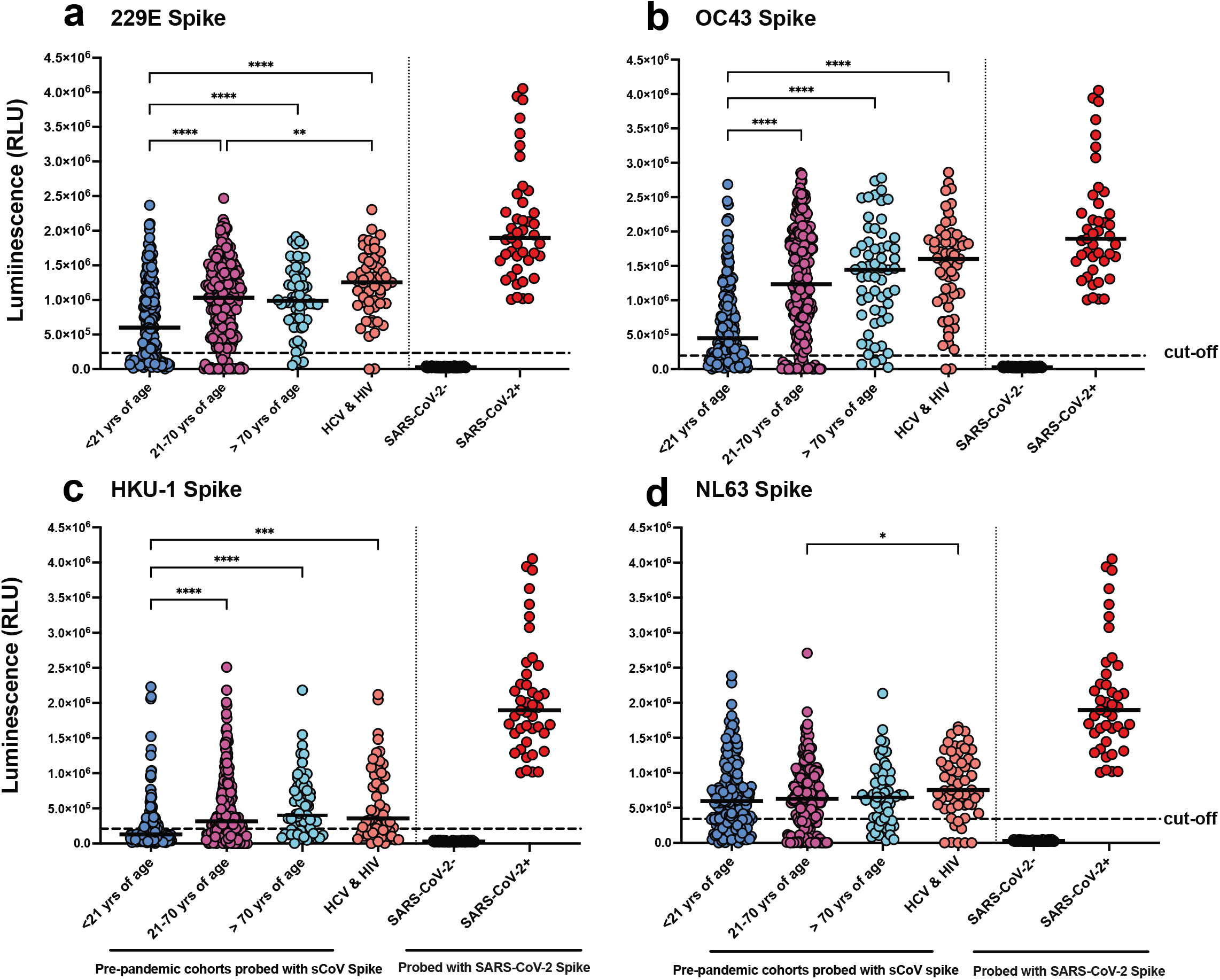
Seroprevalence of human CoVs. ELISA was used to detect spike (S-trimer)-specific IgG antibody responses of four sCoVs: 229E (a), OC43 (b), HKU1 (c) and NL63 (d). Four different cohorts of Pre-COVID-19 pandemic serum and plasma samples were analyzed: < 21 yrs of age [n=193], 21-70 yrs of age [n=251, serum was insufficient to perform all tests on 22 of the entire 273 person cohort], > 70 yrs of age [n=60, serum volume was insufficient to perform all tests on 4 of the entire 64 person cohort], and persons with HCV or HIV [n=50 of which 9 patients were sampled twice at 6 or 12 months intervals for n=59 samples]. Demographics are in Table 1. Note in each panel the same control groups are presented as a range comparison. Controls were SARS-CoV-2 negative individuals from a surveillance study [n=115] and SARS-CoV-2 PCR-confirmed individuals [n=43] challenged against the SARS-CoV-2 S-trimer protein. Demographics of the pandemic cohorts are in Table 1. Dotted lines represent the cut-off values calculated from seronegative pediatric samples (Fig. S1). Statistics were one-way ANOVA with Welch correction and Games Howell *post hoc* multiple comparisons test for pairwise differences. *p<0.05, **p<0.01, ***p<0.001, ****p<0.0001.

### Detection of anti-SARS-CoV-2 Responses in Serum Samples by ELISA

The ELISA procedure is a modified method of a recent published study (16). Antibody responses in pre-pandemic serum samples of all four cohorts were tested for the presence of antibody isotypes (IgA, IgM, IgG, and IgE) and IgG subtypes (IgG1, IgG2, IgG3, and IgG4) using this ELISA binding assay. SARS-CoV-2 antigens S, S-RBD, and N were diluted in sterile 1X PBS (Multicell #311-010-CL) to 2 µg/mL and used to coat a 96-well plates (VWR #62402-959) using 50µL and incubated on a 4°C shaker overnight. The next day, the plates were washed three times with PBS-T and were subsequently blocked with buffer (PBS-T + 3% non-fat milk powder) for one hour. The patient serum samples were diluted 1/50 in dilution buffer [1X PBS + 0.1% Tween (Fisher #BP337-500) (PBS-T), + 1% non-fat milk] in a final volume of 100µL

Accordingly, the control calibration antibodies were prepared with appropriate dilutions to generate the calibration curve (anti-COVID Ig). The dilutions for each curve were as follows: total IgG and IgG1 1/5000 followed by 1:2 serial dilutions; IgM, IgA, and IgG2 1/4000 followed by 1:2 serial dilutions; and IgG3 and IgG4 1/10,000 followed by 1:2 serial dilutions. Blocking solution on ELISA plates was removed after one hour followed by the addition of 100µL of the diluted serum samples and control antibodies to the appropriate wells. The plates were incubated with samples for two hours at room temperature (RT) on a shaker (700rpm). After two hours, plates were washed three times with 200µL PBS-T followed by the addition of secondary-HRP antibodies (1:3000) diluted in dilution buffer. 50µL of the appropriate diluted secondary antibody-HRP was added to each well and the plates were incubated at RT for one hour of shaking (700rpm). After one hour of incubation, the plates were washed four times and developed using OPD tablets (Sigma, P9187) dissolved in 20mL of water for infection (WFI) (Thermo Fisher A1287301). After ten minutes incubating in the dark, 50µl of 3M HCl (Fisher #7647-01) was added to each well to stop the reaction and the optical density (OD) at 490nm was measured using a BIO-TEK Power Wave XS2 Plate Reader. Wells filled with dilution buffer in place of serum represented background controls and were subtracted from the patient serum values.

### Calculation of Cut-Off Values

To establish consistent and unbiased cut-offs, a systematic approach for each combination of antigen and isotype was used. Using the blank subtracted values from the serological assays a preliminary cut off was established at 2 standard deviations (2SD) of the mean of all values. Values over that threshold were then excluded from further calculation of the final cut-off. To establish the final cut-off, the mean and standard deviation were recalculated using exclusively values under the preliminary cut-off. A final cut-off was then established at 2SD of the mean of the distribution under the preliminary cut-off. Values over that final cut-off value were considered positive (ratio to cut-off higher than 1) and values under the final cut-off were considered negative.

To establish the seasonal coronavirus cut-offs, a set of pediatric samples aged between 1-2 years old was used as the reference population due to the high prevalence of seropositivity in adults. The same two pass exclusion strategy at 2SD of the mean was used on that reference group as described above.

### Surrogate Neutralization ELISA (snELISA) assay

To evaluate potential neutralization activity against SARS-CoV-2 S protein, we carried out a surrogate neutralization ELISA based on (25). This neutralization assay was performed using 384-well Immuno plates (Thermofisher, 460372) coated with full length SARS-CoV-2 S-trimer protein diluted in PBS-T (30µL/well of 2µg/mL), followed by an overnight rotating incubation at 4^0^C. The next day, plates were washed two times with 50μl of PBS-T and further incubated with 40µL blocking buffer for one hour on a plate shaker at a speed of 400 rpm. After incubation the plates were washed with PBS-T followed by the addition of pre-COVID-19 serum patient samples diluted 1:4 in dilution buffer (20 μL/well). The plates were further incubated for one hour and washed three times with PBS-T. Biotinylated ACE2 was diluted in dilution buffer to 0.05ng/µL, followed by 20 μL being added to each well (1 ng/well) and a one-hour incubation (400 rpm). After incubation the plates were washed three more times with PBS-T followed by incubation with 20µL of 5ng/µL Streptavidin-Peroxidase polymer diluted in dilution buffer (Sigma#S2438) for 1 hr at 400 rpm. After incubation the plates were washed three times with 60µL of PBS-T. 20µL of SuperSignal ELISA Pico Chemiluminescent substrate diluted 1:2 in PBS was added to each well to measure the luminescence (RLU) signal. Neutralizing activity was determined by calculating the % inhibition against SARS-CoV-2 S proteins using the following equation:

*% Inhibition = 100 – [(average mean of the serum sample/control median)*100]*.

### Statistical and Machine learning analyses

Data analysis was conducted using GraphPad Prism v9, R and Matlab 2021a (Matworks Inc.). Univariate analysis was performed using one-way or two-way ANOVA where appropriate. Welch corrections were applied to all analyses. *Post* hoc tests were Games-Howell’s multiple comparisons when group sizes were greater than 50 or Dunnett T3 when less than 50. Sample clustering was performed using fuzzy c-means (FCM) clustering method (fcm function running under Matlab). FCM allows each feature to belong to more than one group by providing a degree of membership, “belonging”, to each cluster through maximizing proximity between similar features and distance between dissimilar features. FCM is based on the minimization of the objective function: 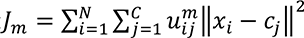 where *m* ∈ (1, ∞) is the “fuzzyfication” factor (with *m*=1 equating FCM to crisp K-means clustering), 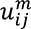 is the membership degree for feature *x_i_* to the cluster *j* with *c_j_* defining the cluster center. Higher membership value indicates stronger belonging to the cluster with membership value of 1 indicating that feature is only associated with the single cluster. Sample clustering was based on serum antibody cross-reactivity to SARS-CoV-2 antigens (N, S-RBD, S-trimer) for IgA, IgM and IgG and capacity of these antibodies to inhibit Spike-ACE2 binding measured for 507 samples. FCM clustering fuzzyfication factor *m* was 1.3 with FCM optimization running until minimum improvement in objective function is 1e^-5^.

Correlation analysis was performed using Pearson and Signed distance correlation metrics calculated on z-score normalized features within each group divided into Responders and combined set of Non-responders and Partial responders. Signed distance correlation was performed using an in-house Matlab routine based on the distance correlation method presented by Szekely and Rizzo (36) with correlation signs derived from Pearson correlation analysis.

Distance correlation *p*-values were calculated using Student’s *t* cumulative distribution function (tcdf function in Matlab). Correlation was determined for 507 samples. Out of this set 12 samples were missing measurements for sCoV. Therefore, correlation calculations were performed both for only 495 samples excluding samples with missing data and for KNN imputed dataset (with N=10 using Euclidean distance). Two approaches obtained the same result. Both Pearson and Distance correlations with p<0.05 and absolute value above 0.2 were shown in the Figure 5. Correlation was performed separately for Responder group (Neutralization value ≥ 26%) and remaining subjects (Neutralization value <26%). Shown are correlation values for the responder group. Group of Non-responders and Partial responders had no correlation for the represented groups at shown threshold levels.

Feature selection was performed using machine learning method Relieff (37). Relieff predicts classification rank, i.e. within the group significance, for each feature in the training set classification, of serum antibody cross-reactivity to 4 different SARS-CoV-2 antigens (N, S-RBD, S-trimer) for IgA, IgM and IgG and separately for antibodies for seasonal viruses (OC43, HKU-1, 229E, NL63). Relieff provides feature selection for classification, i.e. regression, to a continual variable in this case the % inhibition of ACE2-Spike protein interaction. Relieff ranks predictor features by weight for regression to response vector. Negative predictor weight indicates that this is not a good predictor feature and large positive weights are assigned to important predictors. Feature weight decreases if it differs from that of features in nearby instances of the same class more than nearby instances of the other class.

Gaussian process regression (GPR) method was used for the development of a model of percent inhibition of ACE2-Spike protein interaction from the antibody levels for seasonal viruses. GPR is a nonparametric, probabilistic model that introduces latent variables as a function of measured features (here viral vectors) where the new feature function, *f*(*x*), has joint Gaussian distribution (for all features) with optimized kernel that is used to ensure feature distance dependent correlation. Optimized model is: *y = h(x)^T^ β + f(x)* - where *y* is the fitted function (target), *x* are the variables in this case measurement of seasonal virus antibodies, *h*(*x*) are a set of basis functions that transform the original feature vector *x* and β is the vector of basis function coefficients, weights, for contribution of *h*(*x*) to the model. Kernel function used in the model was rational quadratic following optimization. Both feature selection and GPR were performed for 507 samples. GPR provides the regression model with confidence interval as well as predictor weights for the model.

Sample classification was performed using two-layer neural network analysis with 10 hidden neurons and trained with Levenberg-Marquardt backpropagation algorithm with cross-validation with 70% training, 15% test and 15% validation sample division. Classification was performed with sample labels as responsive and non-responsive and using seasonal virus measurements only.

### SARS-CoV-2 ELISA Antigens and Antibodies

To evaluate the functional antibody responses of sCoV antibodies against different SARS-CoV-2 proteins, SARS-CoV-2 S-trimer, S-RBD, and N proteins were used as coating antigen (see antigen production). The following secondary and control antibodies were used during ELISA experiments. Secondary antibodies: Anti-human IgG-HRP (NRC anti-hIgG#5-HRP fusion), anti-human IgA-HRP RRID:AB_2337580 (Jackson ImmunoResearch, 109-035-011), anti-human IgM-HRP RRID:AB_2337588 (Jackson ImmunoResearch 109-035-129), anti-human IgE-HRP RRID:AB_258466 (Sigma A9667-2ML), anti-human IgG1-HRP RRID:AB_2796627 (Southern Biotek 9054-05), anti-human IgG2-HRP RRID:AB_2796633 (Southern Biotek 9060-05), anti-human IgG3-HRP RRID:AB_2796699 (Southern Biotek, 9210-05), and anti-human IgG4-HRP RRID:AB_2796691 (Southern Biotek, 9200-05). Control antibodies: Anti-SARS-CoV-2 S CR3022 Human IgG1 (Absolute Antibody, Ab01680-10.0), Anti-SARS-CoV-2 S CR3022 Human IgA (Absolute Antibody, Ab01680-16.0), Anti-SARS-CoV-2 S CR3022 Human IgM (Absolute Antibody, Ab01680-15.0), Anti-SARS-CoV-2 S CR3022 Human IgE (Absolute Antibody, Ab01680-14.0), Anti-SARS-CoV-2 S CR3022 Human IgG1 (Absolute Antibody Ab01680-10.0), Anti-SARS-CoV-2 S CR3022 Human IgG2 (Absolute Antibody Ab01680-11.0), Anti-SARS-CoV-2 S CR3022 Human IgG3 (Absolute Antibody Ab01680-12.1), and Anti-SARS-CoV-2 S CR3022 Human Ig4 (Absolute Antibody Ab01680-13.12).

### Presence of Reactive IgGs to sCoVs

To determine if the pre-COVID-19-pandemic sera contained IgGs against the four sCoVs, the S-trimer proteins of 229E and NL63 were purchased from creative diagnostics (DAGC134-HCoV-229E; DAGC133-NL63), while OC43 and HKU-1 S-trimer proteins were produced for the study (see Antigen Production below). Using our High Throughput Serological Platform (Hamilton Microlab Star, Biotek 405Ls) we automated the Elisa protocol described in this paper with slight modifications. The serological assay was performed in 384 high binding plates (Thermo Fisher, 460372) using a volume of 12.5 µL to coat each well. Plates were blocked using 80µL of blocking buffer per well. The concentration of antigen, antibodies, and sample diluents were kept as above, although the volume was reduced to 10µL per well. To develop the plates a Luminescent substrate (Thermo Fisher, 37069) was used and RLU measured by a Biotek Neo2 plate reader.

### Antigen Production

To produce the S-RBD of SARS-CoV-2 a plasmid encoding the S-RBD (MN908947) containing amino acid 319-541 with an N-terminal secretory protein sequence was generously given to us by Dr Florian Krammer (Mount Sinai, NYC). The 229E S-RBD (P15423) was cloned in pCAGGs plasmid with a hexa-his Tag (C-term) with secretory signal (N-term) and transfected into 293F cells RRID:CVCL_D603 maintained in Freestyle 293 expression media (Thermo Fisher, 12338018) at 37°C, 7% CO2, with shaking (125rpm). A total of 600 millions cells in 200mL of media were transfected with 200µg of the respective plasmid using ExpiFectamine (Thermo Fisher, 14525). Three days post-transfection the cells were spun down at 4000g for 20min at 4°C and supernatants filtered through a 0.22µm stericup vacuum filter (Millipore Sigma, S2GPU10RE). The filtered supernatant was purified using a Ni-NTA resin (Qiagen, 30210), and washed four times with a washing buffer containing 20nM imidazole. The protein of interest was then eluted with three column volumes of the elution buffer containing 234mM of imidazole. The eluted volume was concentrated and buffer exchanged for PBS using a 10kDa Amicon filter (Millipore Sigma, UFC901008). Concentration was measured and integrity verified by SDS-Page.

The spike ectodomain (SARS2: MN908947; OC43: AAT84362.1; HKU1: Q0ZME7.1) cDNA constructs with furin site mutated, two stabilizing prefusion proline mutations, the human resistin as trimerization partner and a C-terminal FLAG-(His)6 (SARS2) or FLAG-Twin Strep Tag-(His)6 (OC43 and HKU1) tag were cloned into pTT241 vector and transfected in CHO2353 to generate stable pools. SARS-CoV-2 spike was purified by IMAC as described previously while OC43and HKU1 spikes were purified by IMAC followed by StrepTrap HP (Cytiva) affinity chromatography according to the manufacturer’s instructions. All purification steps were performed at room temperature. Integrity and purity of the purified spikes was analyzed by SDS-PAGE and analytical size-exclusion ultra-high performance liquid chromatography (SEC-UPLC).

The nucleocapsid (SARS2: YP_009724397; OC43: AY391777; HKU1: HM034837) cDNAs with a C-terminal FLAG-Twin Strep Tag-(HisG)_6_ tag were synthesized by Genscript (Cricetulus griseus codon bias) and cloned in the cDNA into pTT5™ expression plasmid. Expression was achieved by transient gene expression in CHO^55E1^ cells (38) using polyethylenimine as a transfection reagent. The protein was purified from the clarified cell culture supernatant harvested at day 7 post-transfection. Following centrifugation and filtration, clarified supernatant was loaded on a Nickel Sepharose Excel column (Cytiva Life Sciences). The column was washed with 50 mM sodium phosphate buffer pH 7.0 containing 25 mM imidazole and 300 mM NaCl and protein was eluted with 50 mM sodium phosphate buffer pH 7.5 containing 300 mM imidazole and 300 mM NaCl. Nucleocapsid protein was further purified by affinity chromatography on a StrepTrap™ XT column (Cytiva Life Sciences) equilibrated in 100 mM Tris pH 8.0, 150 mM NaCl (Buffer W). Following washing with 5 column volumes of Buffer W, bound proteins were eluted with Buffer W containing 50 mM biotin and 1 mM EDTA. Purified nucleocapsid protein was buffer exchanged in DPBS using a Centripure P100 column, sterile-filtered through 0.2 µm membrane, aliquoted and stored at −80°C. All purification steps were performed at room temperature. Integrity and purity of the purified nucleocapsid was analyzed by SDS-PAGE and analytical size-exclusion high performance liquid chromatography (SEC-HPLC).

The human ACE2 (Q9BYF1: aa 20-613: TIEE…WSPY) cDNA with an N-terminal human interleukin-10 signal peptide (MHSSALLCCLVLLTGVRA) followed by a Twin-StreptagII – (His)6 – FLAG tag was synthesized by Genscript with codon-optimization for expression in CHO cells. A biotin acceptor peptide sequence (BAP: GLNDIFEAQKIEWHE) was added in-frame at the C-terminus of ACE2. The cDNA was cloned into pTT5 and ACE2-BAP cDNA was expressed by transient gene expression in CHO^55E1^ cells as described above with the addition of 5% (w:w) of pTT5-BirA (E. coli biotin ligase) expression plasmid. Clarified culture supernatant harvested at 8 days post-transfection was purified by IMAC on nickel Sepharose excel as described above. IMAC eluate was loaded on a Strep-Tactin XT Superflow (IBA, Gottingen, Germany), following manufacturer’s instructions. Pooled Strep-Tactin eluate (buffer-exchanged into DPBS) was stored at −80°C.

## Results

### Exposure to all four sCoVs is endemic throughout the population

We acquired human serum and plasma samples, drawn prior to 2019, thus ensuring no previous exposure to SARS-CoV-2. We performed a cross-sectional profiling where a total of 580 patients were grouped into four cohorts based on age and viral infection. Cohorts were: pediatric samples of children and young adults less than 21 years of age (n=193), adults 21-70 yrs of age (n=273), adults greater than 70 years of age (n=64), and persons living with HCV or HIV (n= 50) of whom nine were followed longitudinally (Table 1). Nine patients received two longitudinal blood draws and sCoV seroprevalence was assessed at both time points (separated by 6-12 months) for a total of 589 samples. Each cohort was sex-balanced with mean age equivalence across sexes (Table 1).

We first investigated the prevalence of pre-exposure to the four endemic sCoVs in the pre-COVID-19-pandemic cohorts (Fig.1). Although the four sCoVs (HCoV-OC43, HCoV-229E, HCoV-HKU1, and HCoV-NL63) are prevalent worldwide (39). However, there seems to be a pattern of reinfections by sCoVs that cycles every 12 months (3). We screened pre-COVID-19 serum samples for sCoV anti-S IgG (229E, OC-43, HKU-1, and NL63) using our high-throughput ELISA platform (40, 41). As a reference to frame the dynamic range of our assays, SARS-CoV-2 anti-S data of confirmed negative (pandemic) (n=115) and SARS-CoV-2 PCR-confirmed positive samples (n=43) is presented (Table S1). Our results show that a majority of pre-pandemic samples were seropositive for IgG antibodies against OC43, NL63, and 229E S proteins (Fig. 1 and Table 2).

**Table 2.** Seroprevalence of seasonal coronavirus (OC43, HKU-1, 229E and NL63) IgG antibodies and IgG cross reactivity to SARS-CoV-2 antigens by cohorts. False discovery rate (FDR) was used to describe the percentage of cross reactivity by single and a combination of antigen (S-RBD, N, S-trimer). Combined weighted cross reactivity and seroprevalence is also shown.

| Cohorts | sCoV Prevalence (%) |  |  |  | SARS-CoV-2 IgG FDR (%) |  |  |  |  |  |  |
| --- | --- | --- | --- | --- | --- | --- | --- | --- | --- | --- | --- |
|  | OC43 | HKU-1 | NL63 | 229E | Single antigen |  |  | Two antigens |  |  | Three antigens |
|  |  |  |  |  | RBD | S | N | RBD + S | RBD + N | S + N | RBD + S + N |
| **HCV & HIV** | 96.6 | 72.9 | 86.4 | 96.6 | 8.5 | 6.8 | 15.3 | 1.7 | 3.4 | 1.7 | 0.0 |
| **> 70 Yrs** | 93.3 | 71.7 | 73.3 | 93.3 | 1.6 | 1.6 | 14.1 | 0.0 | 0.0 | 0.0 | 0.0 |
| **21-70 Yrs** | 85.3 | 75.3 | 74.3 | 86.1 | 3.7 | 4.4 | 12.5 | 1.1 | 0.7 | 0.7 | 0.4 |
| **< 21 Yrs** | 72.5 | 34.7 | 70.5 | 68.9 | 5.7 | 6.2 | 6.7 | 2.6 | 2.6 | 1.0 | 1.0 |
| **Weighted Total (all)** | 82.9 | 60.7 | 74.6 | 82.1 | 4.6 | 4.9 | 11.0 | 1.5 | 1.6 | 0.9 | 0.5 |

### Children and adolescents display lower prevalence to sCoVs

One major hurdle in estimating the seroprevalence of IgGs against the four sCoVs is the lack of a true negative reference population. To overcome this challenge, we used a small subset of pediatric samples aged from 1 to 2 years old and established the cut-off at two standard deviations of their mean (Fig. S1). The advantage of using these samples is that it limits the probability that these individuals were exposed to the virus due to their young age, although there is still the possibility of vertically-transferred IgG from the mother through breastmilk (42–44). From these cut-offs, we were able to estimate a relative seroprevalence of sCoVs in the four cohorts. We found that children and adolescents and young adults less than 21 years of age had significantly lower 229E, OC43, and HKU1 sCoV seroprevalence than all other cohorts (Fig. 1 and Table 2). For example, seroprevalence of HKU-1 in persons < 21 yrs of age was about 34.7%, which agrees with Dijkman et al. who found HKU-1 seroconversion in pediatric individuals to be around 36% (45). Additionally, the total cohort weighted HKU-1 average seroprevalence of 60.7% is in agreement with Severance et al. which calculated the seroprevalence of HKU-1 at 59.2% (46). Furthermore, this lower seroprevalence in pediatric samples corroborates previous observations where seroconversion from 229E and NL63 exposure occurs on average 3.5 years following birth (7, 47) (Fig. S1). Across all cohorts, we found an overall size-weighted seroprevalence of 82.9% for OC43, 82.1% for 229E, and 74.6% for NL63 (Table 2). Our calculated seroprevalence for OC43 and 229E is in line with previous studies (5, 46, 48).

### sCoV antibodies are cross-reactive

To ensure the specificity of sCoV detection, we tested co-reactivity between the S-trimer proteins of each of the sCoV along with an additional antigen (S-RBD or N) on a subset of samples (Fig. 2A). It is expected that upon virus exposure and seroconversion, antibodies are detected against both S-trimer and N, and S-trimer and S-RBD antigens of the same virus. Antigen discordance in single-positive samples is likely the result of S-trimer cross-reactivity to other sCoV. We then compared the proportion of samples that were dual-positive for S-trimer and N of OC43 and HKU-1, and S and S-RBD of 229E (Fig. 2b). Positive concordance between the spikes and alternative antigens (i.e., N or S-RBD) for the four cohorts ranged between 67 – 76% (Fig. 2b). These results highlight that there is a relatively high level of cross-reactivity between the S-trimer proteins of sCoVs which compromises assay specificity. Therefore, these data indicate that an accurate determination of sCoVs seroprevalence should measure antibody levels against more than one antigen to minimize the potential of S-trimer protein cross-reactivity.

**Figure 2:**
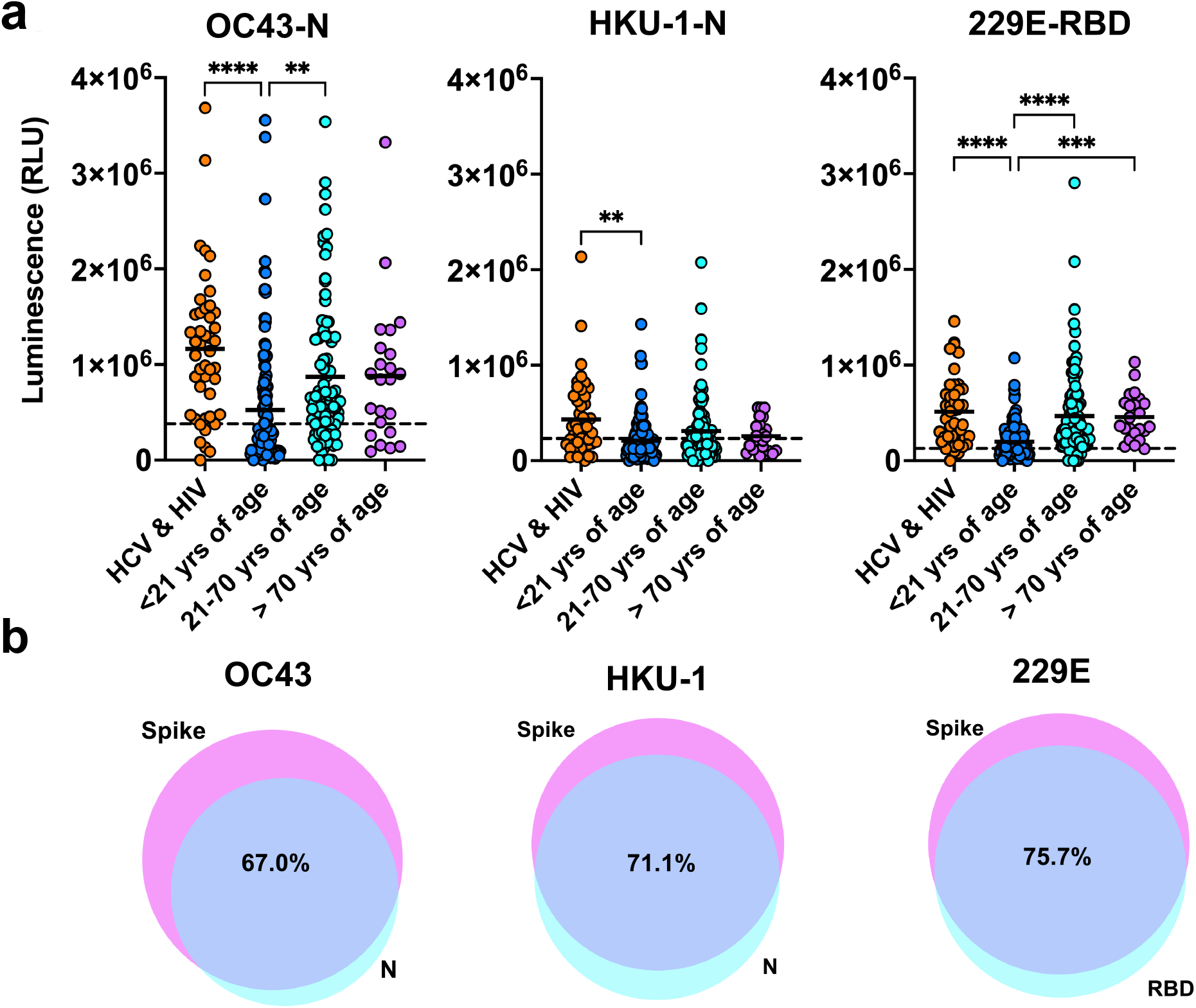
Concordance in sCoV seroprevalence using alternative antigens. (a) Seroprevalence of each cohort < 21 yrs of age [n=129], 21-70 yrs of age [n=111], > 70 yrs of age [n=23], and persons with HCV or HIV [n=45] using alternative antigens is represented (nucleoprotein for OC43 and HKU-1, and S-RBD for 229E). The cut-off was calculated at 2 standard deviation using pediatric samples aged 1-2 years as a negative group. Statistics were one-way ANOVA with Welch correction, *post-hoc* tests were Games-Howell’s multiple comparison tests *p<0.05, **p<001, ***p<0.001, ****p<0.0001. (b) Concordance between using the respective sCoV spike and alternative to evaluate seroprevalence. Percentages of positive concordance between S-trimer and alternative antigens are indicated in each Venn diagram. Venn diagram have been generated using BioVenn online.

### Antibodies to sCoVs cross-react to SARS-CoV-2 antigens

We next assessed the prevalence of IgA, IgM, IgG and IgE binding to SARS-CoV-2 antigens S-trimer, S-RBD, and N in pre-pandemic serum samples (Fig. 3 and Tables 2 and S2-S7) using our high throughput serological assay (31). Cut offs were set at 2 standard deviations (2 SD) above the mean values of the presumed negative sample population. Among pre-pandemic cohorts, 4.6% and 4.9% of samples displayed IgG reactivity against the SARS-CoV-2 S-RBD and S-trimer antigens, respectively (Fig. 3g and 3h, and Table 2). In contrast, the N antigen appeared to be more cross-reactive, showing a higher number of samples that were positive for IgG cross-reactive antibodies (6.7% - 15.3%) (Fig. 3i and Table 2). Given the importance of N as a common target for serological assays (49–51), efforts to accurately measure cross-reactivity to this antigen are critical to account for false positive results. This is especially important when considering that the N antigen of the four sCoVs has on average the highest level of sequence homology to the N of SARS-CoV-2, and that a recent infection by a sCoV could exacerbate such cross-reactivity through immunological imprinting (1, 16). Regardless of their levels, anti-N antibodies are unlikely to offer protection against SARS-CoV-2 entry given that they do not neutralize the virus. Finally, the role of IgE antibodies in mediating immune responses against SARS-CoV-2 infection is currently unknown although IgE antibodies can play a major role in allergic reactions and inflammation (52). Our results show that cross-reactive sCoV IgE antibodies are rare and are unlikely to have a detectable role in influencing SARS-CoV-2 infections (Fig. 3j - 3l, and Table S7).

**Figure 3.**
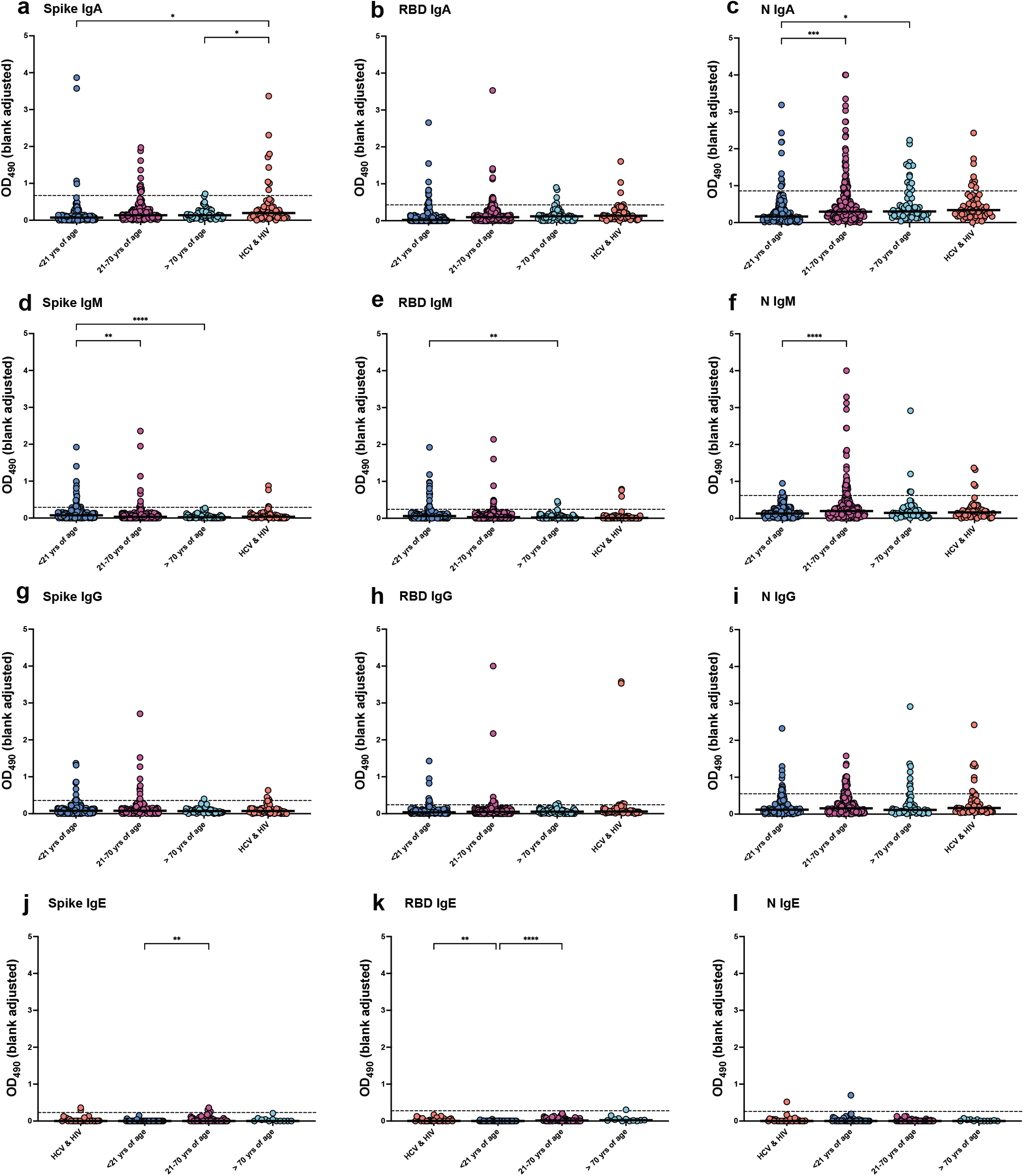
Evaluation of cross-reactive antibodies to three SARS-CoV-2 antigens in pre-pandemic serum samples. We probed for IgA, IgM, IgG and IgE cross-reactive antibodies to the following SARS-CoV-2 antigens: S-trimer, S-RBD and N antigens by ELISA. The levels of IgA (a, b, and c), IgM (d, e, and f), IgG (g, h, and i) and IgE (j, k, and l) are shown as OD_490_ values calculated after adjusting the blank from each serum sample. Dotted lines represent the cut-off values established at 2 SD from the mean value of presumed-negative samples (see Materials and Methods). The false discovery rate (FDR) for each condition are presented in Supplementary Tables S3 to S7. Statistics were one-way ANOVA with Welch correction and Games Howell *post hoc* multiple comparisons test for pairwise differences in A-I where n=193 (<21 yrs of age), n=273 (21-70 yrs of age), n=64 (>70 yrs of age), n=59 (HCV & HIV cohort (50 persons of which 9 were sampled twice 6 or 12 months apart). In panels J-L, serum volumes were sufficient to analyze n=53 (<21 yrs of age), n=68 (21-70 yrs of age), n=12 (>70 yrs of age), n=28 (HCV & HIV cohort). Given sample sizes, statistics were one-way ANOVA with Welch correction and post-hoc tests were Dunnett T3 multiple comparison tests, *p<0.05, **p<001, ***p<0.001, ****p<0.0001.

**Figure 4.**
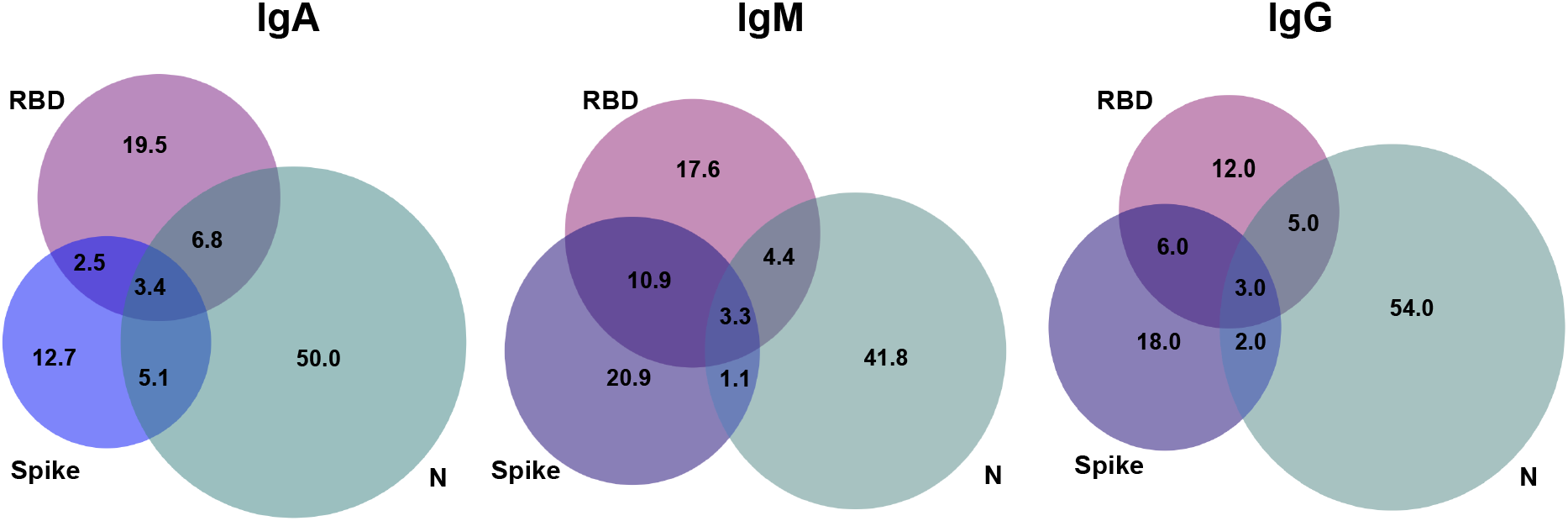
Patterns of multi-antigen cross-reactivity against SARS-CoV-2 Spike-ACE2 interactions. Venn Diagram showing the cross-reactivity patterns across all three antigens probed for IgA, IgM and IgG. Most cross-reactive events are for a single antigen. BioVenn online was used to generate the quantitative Venn diagram (49).

We further investigated cross-reactivity of the IgG1, IgG2, IgG3, and IgG4 isotype subclasses against the three SARS-CoV-2 antigens (Figs. S2, S3, and Tables S2-S7). Our analysis revealed that most of the IgG subclass antibodies in pre-pandemic sera only exhibited low levels of cross-reactivity with SARS-CoV-2 antigens. Detection of cross-reactive IgG3 antibodies is interesting, as it has been shown that SARS-CoV-2 S-specific IgG3 subclass levels increase with COVID-19 disease severity (53). We further wanted to assess the false discovery rate (FDR) when two or three antigens are used for the analysis. For this, we performed a comparison of all the serology results across three antibody isotypes and three antigens to determine if a positive sample is positive for only one or multiple antigens and/or isotypes (Figs. 5, S3 and Tables 2 and S7). We found that in most cases, a positive sample is restricted to only one isotype and/or one viral antigen. This highlights the importance and value of considering simultaneously more than one antigen for SARS-CoV-2 serological testing. We calculated the FDR for each specific cohort disaggregated by individual antigen-isotype/subtype (Tables S2-S6), and we also calculated the FDR for each combination of antigen and isotype/subtype (Tables 1 and S7). While the IgG FDR for our complete pre-pandemic cohort ranged from 5% to 11% when probing individual antigens, these values dropped to between 0.9% to 1.6% when probing two antigens, and 0.5% when all three antigens are used to determine seropositivity (Table 2).

**Figure 5.**
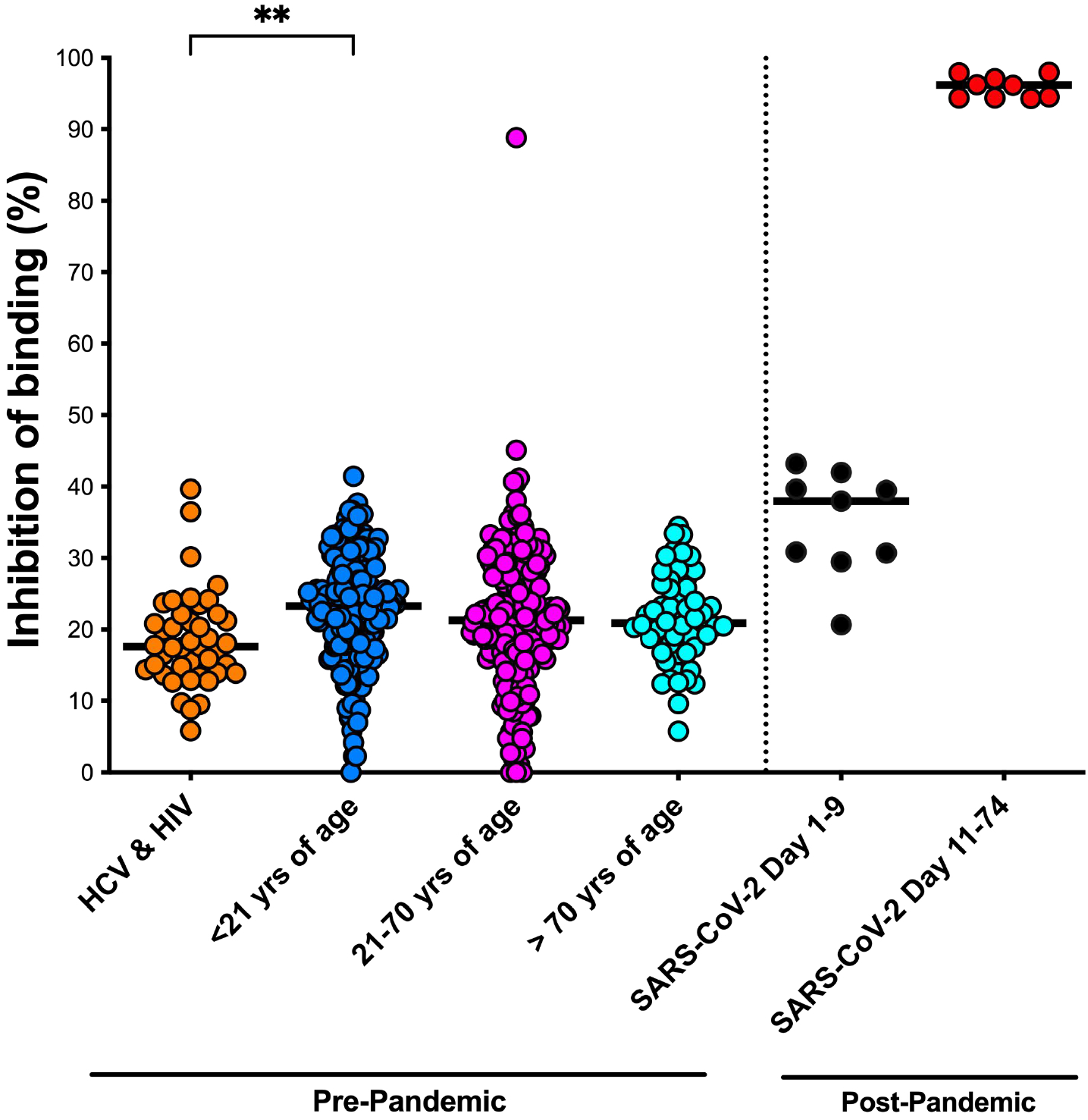
Neutralizing activity against SARS-CoV-2 Spike-ACE2 interactions. Neutralizing activity of 507 serum samples across the four cohorts were tested for the ability to inhibit Spike-ACE2 interactions. The snELISA assay involves coating plates with the SARS-CoV-2 S-trimer protein and applying serum, washing and challenging with biotinylated ACE2 (21). The plot shows the percentage of inhibition relative to the no serum control. Each point is the mean of quadruplicate values. Control samples are constituted of the serum of SARS-CoV-2 infected individuals collected 1 to 9 days after testing positive by PCR (black), or 11 to 74 days post-PCR (red). Statistics were one-way ANOVA with Welch correction, *post-hoc* tests were Games-Howell’s multiple comparison tests *p<0.05, **p<001, ***p<0.001, ****p<0.0001.

### sCoV antibodies partially cross-neutralize SARS-CoV2 antigens

Previous studies have reported the presence of cross-reactive antibodies to SARS-CoV-2 S-trimer protein in pre-pandemic blood samples (1, 12, 28, 54, 55). However, while generally limited information is available pertaining to levels of cross-neutralization of SARS-CoV-2, some information has been published with regards to the neutralization of S-pseudotyped viruses (1, 31). Given the importance of neutralizing antibodies that target the SARS-CoV-2 S-trimer (28, 56–58), we evaluated the relative efficacy of sCoV antibodies to inhibit SARS-CoV-2 Spike-ACE2 interactions using a protein-based surrogate neutralization ELISA (snELISA) assay (54). Inhibition of Spike-ACE2 interaction varied from 0-45%, with a single patient showing inhibition level of 89%, with median levels ranging between 18-23% (Fig. 5, Table 3). Very weak neutralizing activity was previously observed by others in a different experimental system that measured infection by S-pseudotyped viruses (1, 31). By contrast, we identified a moderate yet distinct pattern of neutralization between cohorts of pre-pandemic samples.

**Table 3.** Descriptive statistics of the protein-based neutralization assay by cohorts. Median values and range (minimum and maximum values) of the percentages of the inhibition of SARS-CoV-2 spike-ACE2 was calculated by cohorts from the protein based surrogate neutralization assay (snELISA).

| ACE2 Binding Inhibitions (%) |  |  |  |
| --- | --- | --- | --- |
| Cohort | Median | Min value | Max value |
| HCV & HIV | 17.6 | 5.83 | 39.7 |
| < 21 yrs | 23.3 | 0.00 | 41.4 |
| 21 - 70 yrs | 21.3 | 0.00 | 45.1* |
| > 70 yrs | 21.9 | 5.77 | 34.4 |
| SARS-CoV-2 Day 1-9 | 37.9 | 20.7 | 43.2 |
| SARS-CoV-2 Day 11-74 | 96.2 | 94.3 | 98.0 |

\*Patient P1-G5 was excluded from this analysis (89% inhibition).

### Abundance of sCoV antibodies does not predict SARS-CoV-2 neutralization

Next, we asked if there was an association between sCoV antibody levels with the levels of neutralization observed. Using fuzzy c-means unsupervised clustering, we clustered samples into 10 distinct groups of human sera from 507 of our 580 patients from which we had sufficient material to monitor the degree of inhibition of SARS-CoV-2 Spike-ACE2 interaction and cross-reactivity with SARS-CoV-2 S-RBD, S-trimer, and N proteins (Fig 6a). Subjects stratified independently of age and sex or other prior viral infection (HIV & HCV) (Fig. 6a, Table S8). We found that these clusters defined a continuum of SARS-CoV-2 neutralization in patients ranging from 0% inhibition to 45% (Fig. 6b). Interestingly, this serum sample also displayed high IgG cross-reactivity against S-RBD (O.D.= 2.172) and S-trimer (O.D.=2.707) along with high IgA cross-reactivity against S-RBD (O.D.=0.275) and S-trimer (O.D.=0.233). Additionally, the presence of autoantibodies against ACE2 that block spike binding could also theoretically contribute to the neutralization given that autoantibodies are detected in epilepsy (42–44). Additional neutralization assays using S-pseudotyped viruses are on-going to confirm this observation.

**Figure 6.**
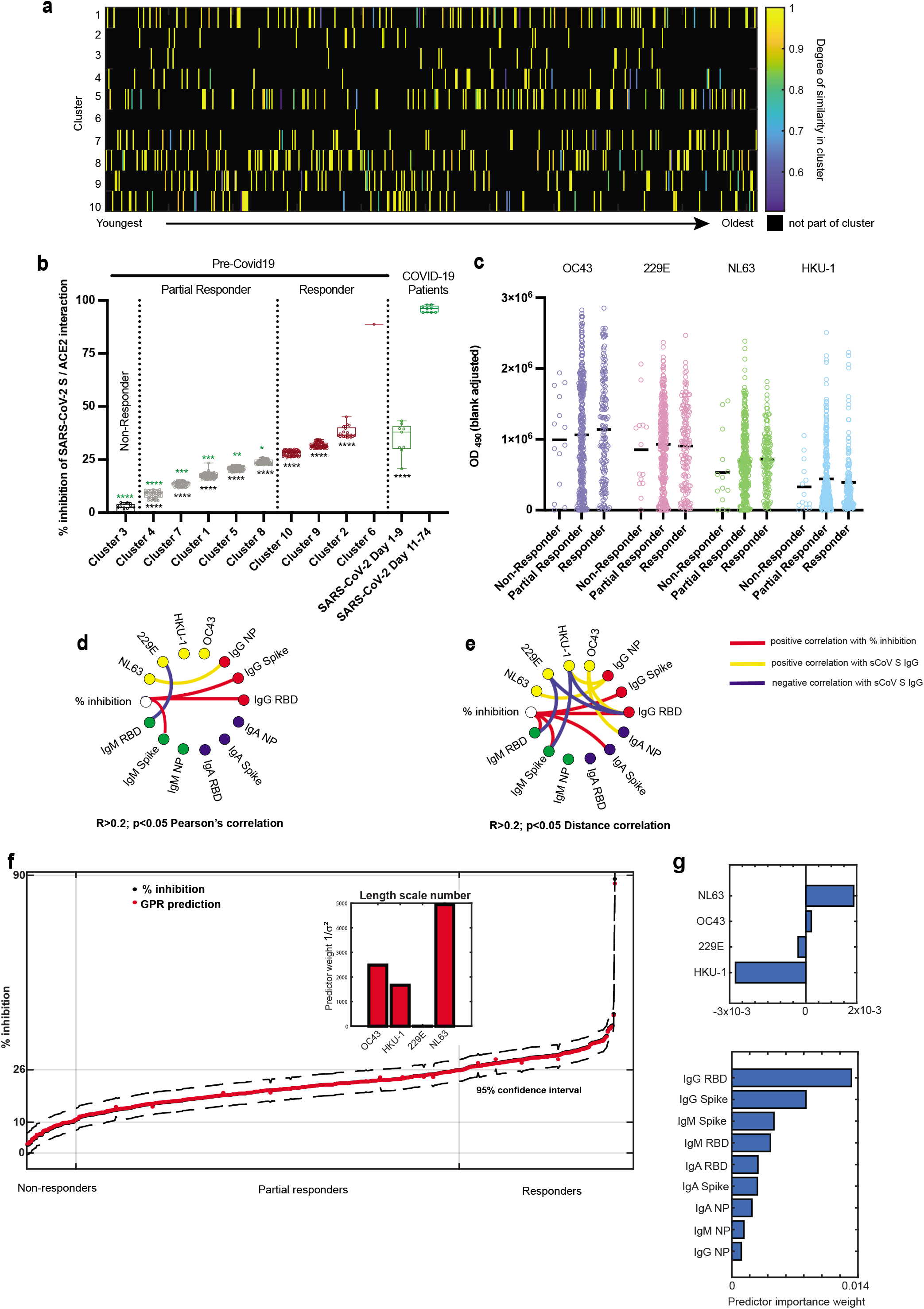
Association of sCoV seroprevalence with SARS-CoV-2 antigen cross-reactivity and neutralization of SARS-CoV-2 Spike-ACE2 interactions. (a) Fuzzy c-means clustering segregated 507 patients into 10 clusters across all 4 cohorts (with fuzzyfication factor m=1.3) based on serum antibody cross-reactivity and capacity of these antibodies to inhibit SARS CoV-2 Spike binding to ACE2. Patient stratification was independent of age or sex (Table S8). Data represent degree of cluster identification as a measure of similarity of all features per patient. Colours indicate membership values over 0.5 - 1.0 with highest values indicating the strongest cluster confidence assignment. (b) Clusters stratified patients with increasing capacity to inhibit interaction between the SARS-CoV-2 spike protein and ACE2. Data represent percentage of inhibition relative to the no serum control. Boxes extend from 25th to 75th percentile. Whiskers delineate minimum and maximum points. Each patient point is the mean of quadruplicate values. Each cluster (with the exception of cluster 6 composed of a single patient with robust neutralizing activity) was compared to control samples constituting of serum of infected individuals collected 1 to 9 days after testing positive by PCR for SARS-CoV-2 (green asterisks) or to Non-Responders (cluster 8, black asterisks). Statistics were one-way ANOVA with Welch correction for unequal standard deviation, post-hoc Dunnett’s T3 *p<0.05, **p<0.01, ***p<0.001, ****p<0.0001 green vs SARS-CoV-2, black vs cluster 3. Subjects that failed to inhibit SARS-CoV-2 Spike-ACE2 interactions were dubbed Non-Responders (black, Cluster 3). Subjects that exhibited significantly higher neutralizing activity than Non-Responders yet significantly lower neutralizing activity than COVID-19 patients (green) were dubbed Partial Responders (grey). Patients that exhibited significantly higher neutralizing activity than Non-Responders yet comparable activity to COVID-19 patients were dubbed Responders (red). (c) Abundance of sCOVs does not discriminate capacity to neutralize SARS-CoV-2 Spike-ACE-2 interaction. There is no statistical difference between sCOV abundances between groups. Statistics were one-way ANOVA with Welch correction for unequal standard deviation, post-hoc Dunnett’s T3. (d) Using Pearson’s correlation level over 0.2 and p<0.05, % inhibition of SARS-CoV-2 spike-ACE2 interaction associated strongly with cross-reactivity with SARS-CoV-2 IgG S-RBD and IgG S-trimer while the abundance of antibodies to NL63 S-trimer correlated with IgG N. The 229-E abundance of antibodies showed negative correlation with IgM S-RBD. (e) Signed distance correlation for Responder group (shown for correlation greater then 0.2, p<0.05) reveal that % inhibition of SARS-CoV-2 spike-ACE2 interaction associates strongly with SARS-CoV-2 S-trimer IgM S-RBD, IgM S-trimer, IgA S-trimer, IgG S-RBD, and IgG S-trimer. Abundance of antibodies to NL63 spike and HKU-1 correlate positively with IgG N, OC43 spike with IgG S-RBD and IgA N. 229E shows negative correlation with IgM S-RBD, IgM S-trimer while HKU-1 shows negative correlation with IgM S-trimer and IgG S-RBD. (f) Gaussian process regression (GPR) model for % inhibition of ACE2-Spike protein interaction predicted from sCoVs antibody abundance showing predictive model with model 95% confidence intervals (dashed lines) as well as predictor weight, i.e., relevance of each feature for the model performance, shown as an inset. In the figure, the importance of the feature increases with the predictor weight shown with NL63 being the most important predictor of latent variables providing GPR model and 229E the least relevant in the model building (g) Relieff feature selection for % inhibition of ACE2-Spike protein interaction determined for sCoVs antibody abundance and SARS-CoV-2 cross-reactivity measures separately. Shown are features’ weight values with negative weight indicating no significance in regression and significance in the model increasing with the predictor importance

We then further sub-divided these patient clusters into three groups: Non-Responders, Partial Responders, and Responders based on their similarity to newly infected SARS CoV-2 patients (Day 1-9 from testing positive) with respect to SARS-CoV-2 neutralization (Table S1). Sera from 3% of all subjects (N=16, Cluster 3) did not neutralize binding of SARS CoV-2 S to ACE2 (Non-Responders, Fig. 6b). Sixty eight percent of all subjects were Partial Responders, exhibiting significantly higher neutralizing activity than Non-responders, yet significantly lower neutralizing activity than newly infected COVID-19 patients (Fig. 6b). Twenty-nine percent of subjects were Responders, exhibiting significantly higher neutralizing activity than Non-responders and comparable activity to that of newly infected COVID-19 patients (Fig. 6b). We asked what pattern of sCoV reactivity against their respective S-trimer proteins discriminated these groups. We found no statistical difference in the levels of antibodies against OC43-S, 229E-S, HKU-1-S, or NL63-S (Fig. 6c). Moreover, the vast majority of cross-reactive SARS-CoV-2 antibodies were directed against the N antigen (Tables 2 and S7). These data suggested that absolute abundance of sCoV anti-S IgG antibodies did not dictate neutralization.

### Relative ratios of sCoV antibodies predict neutralization

To identify critical features underlying neutralization response, we used Pearson’s correlation analysis to determine major linear correlations as well as signed distance correlation to obtain information about linear, non-linear and indirect, and more distant associations. Through these analyses, we were able to discern a pattern of seroprevalence to sCoVs that closely associated with the capacity to neutralize Spike-ACE2 interactions. First, the percent inhibition of Spike-ACE2 binding in the Responder group positively associated with higher levels of SARS-CoV-2 cross-reactive IgG S-trimer and IgG S-RBD antibodies (at a Pearson’s correlation above 0.2 and p<0.05), as expected (Fig. 6d), and additionally with abundances of IgM S-RBD, IgM S-trimer and IgA S-trimer antibodies (at a distance correlation level above 0.2 and p<0.05) (Fig. 6e). Of these five neutralization correlates, abundances of OC43 S-trimer IgG associated with higher levels of SARS-CoV-2 cross-reactive IgG S-RBD and IgA N. Additionally, levels of NL63 S-trimer and HKU-1 S-trimer IgG positively associated with levels of cross-reactive IgG N (Fig. 6e). At lower distance correlation level of 0.17 there was also direct correlation between NL63 and the percent inhibition (not shown). Conversely, abundances of anti-229E S-trimer IgG negatively correlated with both SARS-CoV-2 IgM S-RBD (Pearson’s correlation, distance correlation, Fig. 6d) and IgG S-RBD (distance correlation, Fig. 6e). Finally, HKU-1 S-trimer IgG levels negatively correlated (via distance correlation) with IgM S-trimer and IgG S-RBD (Fig. 6e).

Taken together, these data suggest that NL63 S and, to a lesser extent, OC43 S antibody levels positively associate with correlates of percent inhibition and 229E and HKU-1 negatively associate with correlates of percent inhibition. These data are consistent with Selva et al. who have shown that elderly individuals with higher sCoV exposure show increased levels of cross-reactive SARS-CoV-2 IgA and IgG responses, while children show elevated SARS-CoV-2 IgM and is associated with less frequent exposures (59). Here, we show that these responses link to OC43 S-trimer abundance. In our analysis, cross-reactive SARS-CoV-2 IgM levels negatively correlated with 229E and HKU-1 anti-S antibodies. Thus, a higher abundance of these antibodies is associated with a decreased neutralizing probability following exposure to these two sCoVs according to our model. Percent inhibition is in turn correlated with anti-SARS-CoV-2 IgM (IgM S-RBD and IgM Spike), IgA Spike and IgG (IgG S-trimer and IgG S-RBD), possibly indicating a different source of protection in our responder group members belonging to different age groups. These results are in excellent agreement with the further statistical analysis of the feature significance for classification, regression to percent neutralization determined using Relieff. Although there was no statistically significant difference in the levels of antibodies against sCoVs between the three groups (Fig. 6c), Relieff selection of the most significantly different features related to the continual value of percent neutralization demonstrated that levels of NL63 and OC43 have the positive, i.e., significant, classification weight to percent neutralization (Fig. 6g). This result is supported by their positive correlation with IgG N, IgG S-RBD and IgA N. Finally, IgG S-RBD, IgG S-trimer, IgM S-trimer and IgM S-RBD have the highest classification weight according to Relieff analysis, in agreement with the correlation analysis in Figures 6d and 6e providing additional confirmation that the pattern of sCOV S antibodies in Responders underlies neutralization capacity.

### Cross-reactive antibodies and latent variables impact neutralization

To test this hypothesis computationally, we asked whether we could use only sCoV S reactivity to predict neutralization of the SARS-CoV-2 Spike-ACE2 interaction (Fig. 6f). We used a Gaussian process regression (GPR) method to explore the functional relationship between sCoV S antibody levels and the percent inhibition of ACE2. The GPR model, using sCoV antibody levels as predictors, provided excellent agreement with percent inhibition of ACE2 for all three patient groups (Non-responders, Partial Responders and Responders, Fig. 6g). Because GPR optimizes variable projection, weights, as well as covariance as a function of the latent variables, these results indicate that sCoV antibody measurements, when combined with latent variables, (i.e., variables that are not measured but can be determined as functions of measured values), dramatically improve prediction of percent inhibition of ACE2 interaction. GPR analysis also provides a confidence interval of this prediction, i.e., error level of the prediction (indicated in dashed lines in Fig. 6f). The significance of a variable in the developed GPR model, measured as the weight of the predictor indicates major significance of NL63 in modeling followed by OC43 and HKU-1. In this analysis predictor weight of 229E is significantly lower. This is once again in agreement with the correlation analyses demonstrating 229E is indirectly negatively correlated with percent inhibition (Fig. 6e). Global sensitivity analysis (GSA) apportioning the outputs uncertainty to the uncertainty of each input factor over their entire range and with input factors varied simultaneously is calculated using Sobol method and quasi-random Monte Carlo sampling. GSA shows once again major significance of NL63 in the variance of the output and second highest uncertainly contribution of 229E, showing in correlation analysis major negative correlation with the % neutralization. GSA result for GPR model presented in Figure 6 is shown in Supplementary Figure 4. Taken together, these data link the capacity of pre-pandemic sera to neutralize SARS CoV-2 Spike-ACE2 binding to both specific patterns of elevated antibody levels against NL63 S-trimer and OC43 S-trimer as well as additional unknown factors, yet to be identified, conferred by this specific pattern of cross-reactive antibodies to SARS-CoV-2 S (Fig. 6e).

## Discussion

Overall, our data has highlighted that there is a very large range in antibody levels against sCoVs. Interestingly, measuring seroprevalence of sCoVs using a single antigen is prone to specificity challenges due to cross-reactivity between sCoV antigens. We show that only 67% to 76% of spike-positive samples were also positive when probed with a second antigen from the same virus (Fig. 2b). This indicates that we are over-reporting seroprevalence of individual sCoVs. Additionally, while there is no statistical difference in prevalence between adults and elderly individuals, there appears to be a modest decrease in prevalence in younger individuals for 229E, OC43 and HKU-1. A recent paper by Selva et al. also observed that children have fewer exposures to sCoVs than older adults (59). Exactly how high levels of antibodies against sCoVs or recency of infection by sCoVs may influence COVID-19 disease severity remains unclear. However, our data highlights relatively high levels of cross-reactivity against certain SARS-CoV-2 antigens, with N being the most frequently detected at 11% overall prevalence, while S-trimer displayed about 5% prevalence (Table 2). These values are comparable to another study that showed 16.2% cross reactivity to N and 4.2% for S-trimer (1). Small differences between our two studies can be accounted for by cohort composition that affects certain antigen/isotype pairs disproportionately (Figs. 3, S2 and tables S2-S6). Also, differences in S-RBD and S-trimer FDR (Table 2), and thereby assay specificity, can be caused for by the methods used for antigen production. Glycosylation patterns can vary depending on the cell expression system and also, S-RBD can potentially expose new epitopes that might be hidden when it is part of the full S-trimer.

Although some studies have indicated that no detectable SARS-CoV-2 neutralization activity was detected in pre-pandemic blood samples using various types of assays (1, 31, 59), the snELISA that we used was able to discern small variations from 0% to 45% in Spike-ACE2 binding inhibition. Although snELISAs are not capable of providing the same complete assessment of whole virus entry neutralization as measured when using live replicative virus, this methodology does enable an isolated and focused assessment of one parameter of the neutralization which is the binding of the viral spike to its host receptor. Small variations at this level can have profound implications during an actual infection in a human host that could perhaps be missed using cell lines to assess neutralization with live virus, the current gold standard. Spike-pseudotyped lentiviruses are also useful surrogates to study neutralization and viral entry, but lentiviruses bud from the cell surface and likely harbor a different lipid and host molecule compositions in their envelope than SARS-CoV-2 that mostly egress through exocytosis and possibly also through the lysosomal pathways (60, 61). Given these differences in the biochemical and immunological composition of the envelopes of lentiviruses and betacoronaviruses, neutralization data obtained with pseudotyped lentiviruses should not be considered as definitive. Nevertheless, in agreement with previous studies, we also did not identify a direct link between the abundance of sCoV S-trimer antibodies and neutralization of Spike-ACE2 interactions (Fig. 6c). However, our data do indicate that cross-reactive IgG against S-trimer and S-RBD of SARS-CoV-2 have a positive correlation with neutralization efficiency, and also to a lesser degree, IgM against S-trimer and S-RBD, and IgA against S-trimer (Fig. 6d and 6e). An even more interesting observation is how NL63 correlates with cross-reactive N IgGs (Fig. 6d), a similar observation was also made by another group (32). While N antibodies are unlikely to be neutralizing, Fc effector function such as antibody dependant cell mediated cytotoxicity (ADCC), complement activation, their role in CD8+ T cell responses and /or the cytokines and immune factors involved in their production may all have an indirect influence on other antiviral pathways in a live infected host.

Despite a small number of studies reporting undetectable cross-neutralization or unlikely protection by sCoVs antibodies (1, 31, 59), there is a large body of evidence that supports a role for prior exposure to sCoVs with milder COVID-19 disease severity and/or detectable immunity to SARS-CoV-2 (6, 7, 10, 13, 29, 33, 34, 62, 63). Using machine learning approaches, we present here data that reconcile these differences by providing evidence that it is the relative pattern of prior sCoV antibody levels in sera, and not total levels, that predict neutralization intensity levels. Additionally, GPR analysis shows that latent variables, that have not been identified in this work but that can be inferred from the measures of sCoVs, provide a model that accurately predicts the percent inhibition of Spike-ACE2 binding.

In light of the ongoing COVID-19 pandemic, public health measures are geared towards reducing exposure to the virus, reducing mortality and hospitalization. The predictive value of this model offers an alternative way to evaluate neutralization by using serological assays against sCoV’s which are easier, more accessible and use less resources than traditional neutralization assays. Furthermore, our predictive model contributes to the growing body of evidence in support of the protective outcome of prior exposure to sCoVs, NL63 in particular. It is well understood that humoral responses are only one arm of the immune system, evidence that cross-reactive T cells also play a role in protection is also gaining momentum (33). With latent factors being here identified to exert an influence over S/ACE2 binding, an understanding of how sCoV may diminish the severity of COVID-19 will require a broader investigation. Current study design does not allow analysis of causal relationships in vivo and also causal relationship between sCoV and any latent factors. Additionally, further molecular, omics, measurements in these patient samples would be greatly beneficial for understanding of the observed effects. In future work, it will be critical to evaluate if our GPR analysis holds true in predicting COVID-19 severity by way of sCoV antibody ratios.

## Data Availability

All the study raw data will be made publicly available upon request.

## Contributors

YG, VS, EM, GL carried out the experiments. MG and PMG supported assay development. M-AL & YG designed the study. YG, VS, SALB, MCC and M-AL wrote the manuscript. YD contributed to the design of spike, N, ACE2 constructs, expression and purification procedures. RAB sourced specimens, co-designed high-throughput analysis. MM and CC provided samples and AMC facilitated serum sample processing. SALB and MCC contributed to sample collection, data verification / processing and performed all of the sCOV analyses with YG and M-AL. All of the authors have edited and contributed to the review of the manuscript.

## Acknowledgments

The authors would like to thank Anne-Claude Gingras for helpful comments on the manuscript. This study was supported in part by a COVID-19 Rapid Response grant by the Canadian Institutes of Health Research (CIHR; #VR2 - 172722) and by a grant supplement by the COVID-19 Immunity Task Force (CITF) to M-A Langlois and by the National Research Council of Canada Collaborative R&D Initiative Pandemic Response Challenge Program Grant to SAL Bennett and M Cuperlovic-Culf (PR031-1). Production of COVID-19 reagents was financially supported by the NRC’s Pandemic Response Challenge Program. M.-A.L. holds a Canada Research Chair in Molecular Virology and Intrinsic Immunity. S.A.L.B holds a University Research Chair in Neurolipidomics. Y.G. holds a CIHR Frederick Banting and Charles Best graduate scholarship (CGS-M).

## Data sharing statement

The data used in this study can be obtained by request to M.A.L

## Supplementary materials

Supplementary material associated with this article can be found, in the online version, at doi:

## SUPPLEMENTARY MATERIALS

**Supplementary Figure 1.**
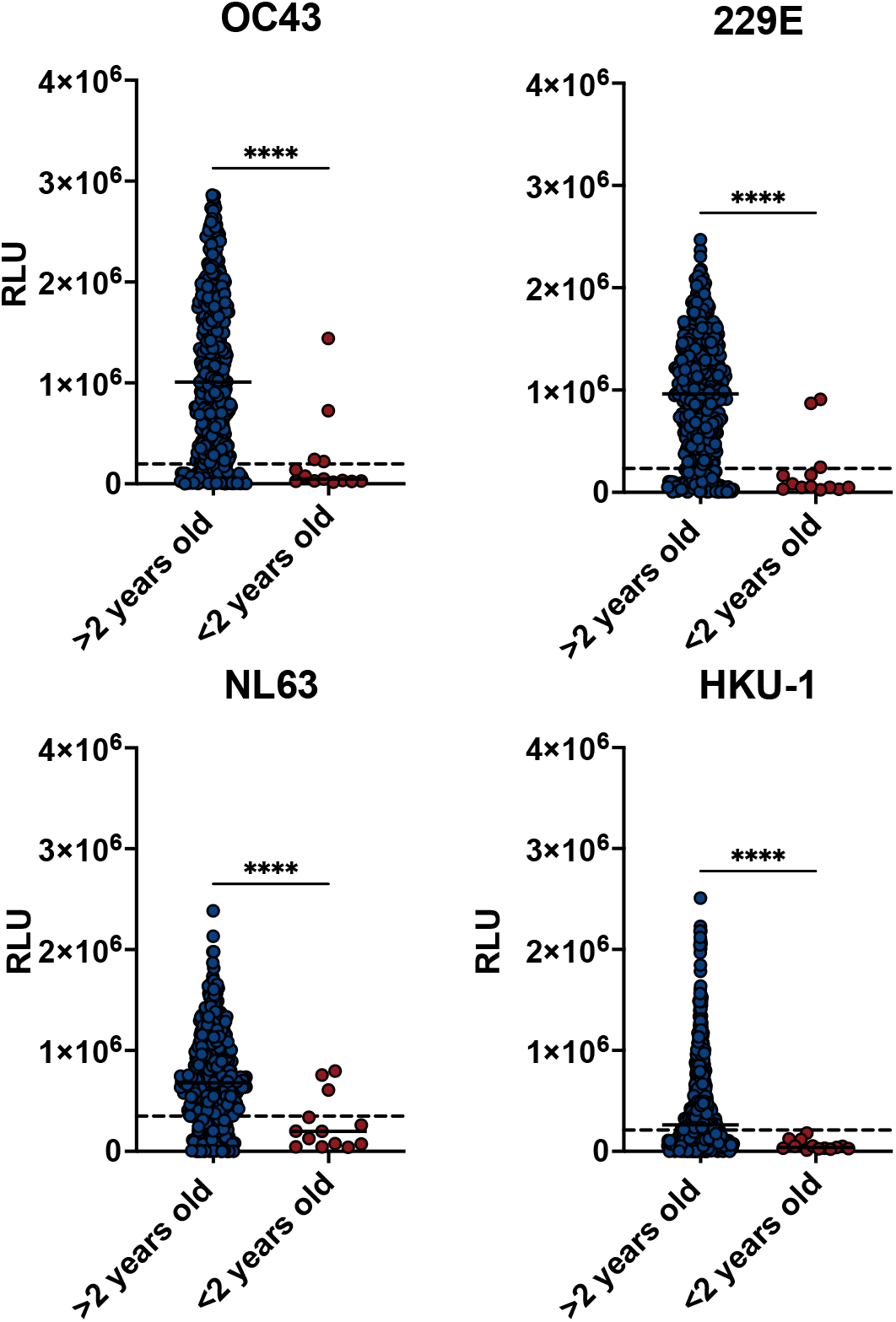
Comparison between pediatric samples under 2 years old to the complete cohort for each seasonal spike coronavirus. Pediatric individuals under two years old that were used to establish the cut-offs are indicated (red) in contrast to the seroprevalence of the combination of the four cohorts (Blue). <2 yrs of age [n=13], >2 yrs of age [n=550]. Welch’s unpaired T test was performed to establish statistical significance *p<0.05, **p<0.01, ***p<0.001, ****p<0.0001.

**Supplementary Figure 2.**
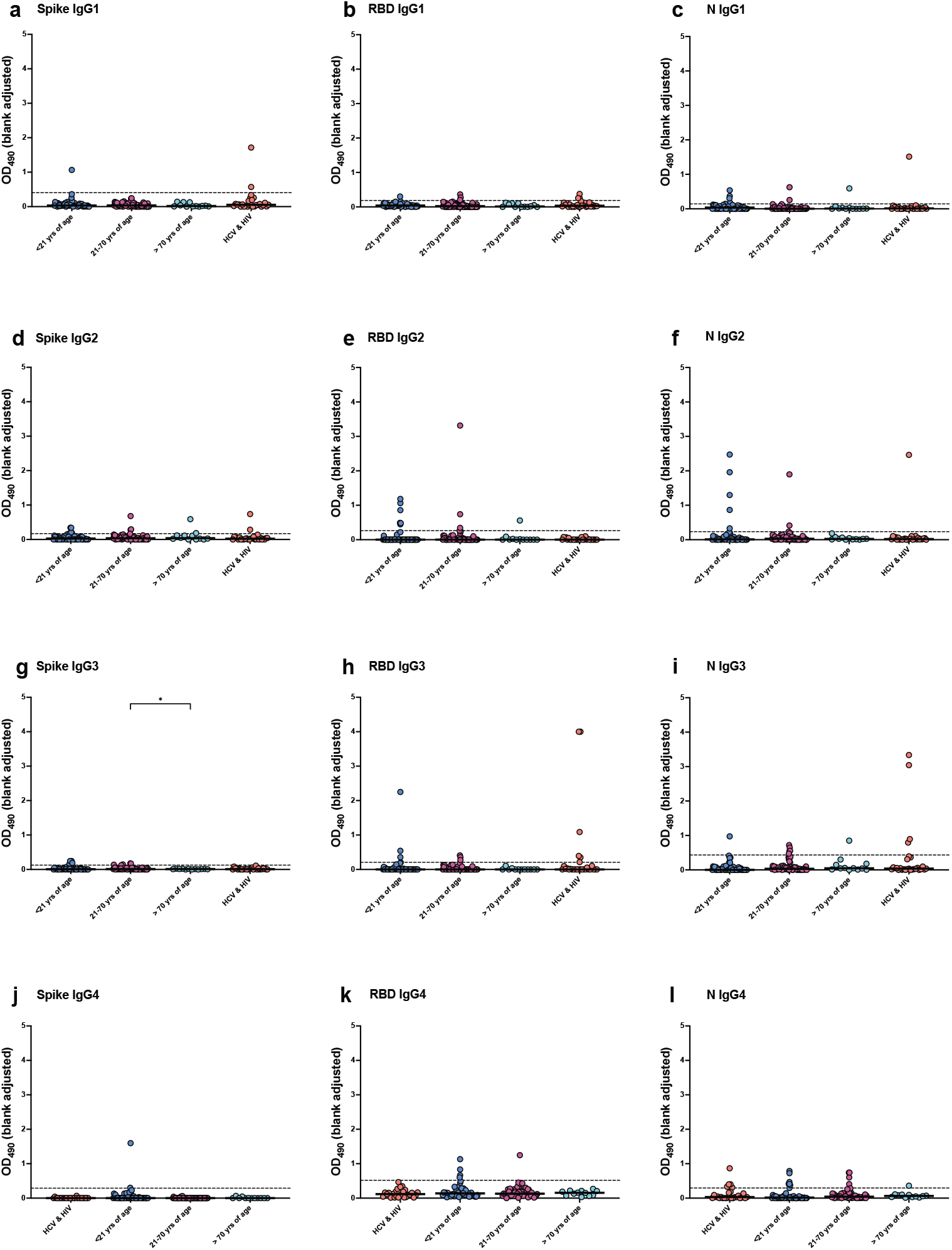
IgG-subclass antibody responses to SARS-CoV-2 antigens. The pre-COVID-19 patient cohort samples (x-axis) of HCV & HIV [n=28]; **< 21 yrs**, n=[53]; **21 - 70 yrs** [n=68]; **> 70 yrs** [n=12]; were tested for the reactivity of IgG1 IgG2 IgG3 and IgG4 to the SARS-CoV-2 S-trimer protein **(a, d, g, j)** S-RBD **(b, e, h, k)** and nucleocapsid **(c, f, i, l)**. The ELISA OD_490_ values indicates the raw OD subtracted from the background. Cut-off values calculated by two rounds of exclusion at 2 Standard deviation from the negative distribution was indicated on each graph. Statistics were one-way ANOVA with Welch correction, *post-hoc* tests were Games-Howell’s multiple comparison tests *p<0.05, **p<001, ***p<0.001, ****p<0.0001.

**Supplementary Figure 3.**
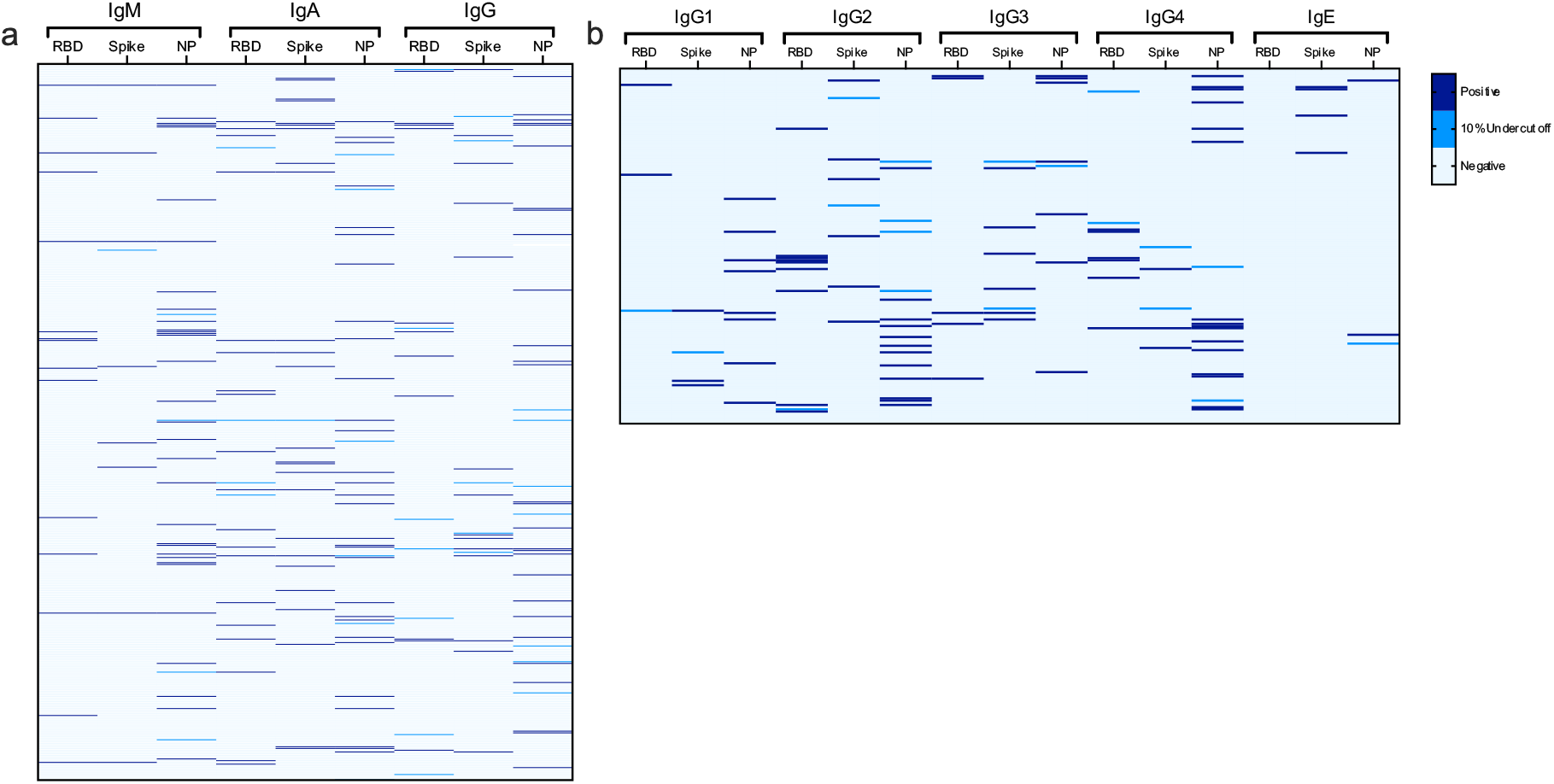
Discordance of serological false positive per isotype and antigen. (a) An alignment of the signal to cut-off ratio for each sample was performed by isotype (IgG, IgM, IgA) and antigens (S-RBD, S-Trimer, N) on 589 samples from which 9 were longitudinal. Values over cut-offs (dark blue) and 10% under cut offs (light blue) are indicated. (b) An alignment of the signal to cut-off ratio for was performed by subtypes (IgG1, IgG2, IgG3, IgG4) and IgE and antigens (RBD, Spike, N) on 161 randomly selected samples.

**Supplementary Figure 4.**
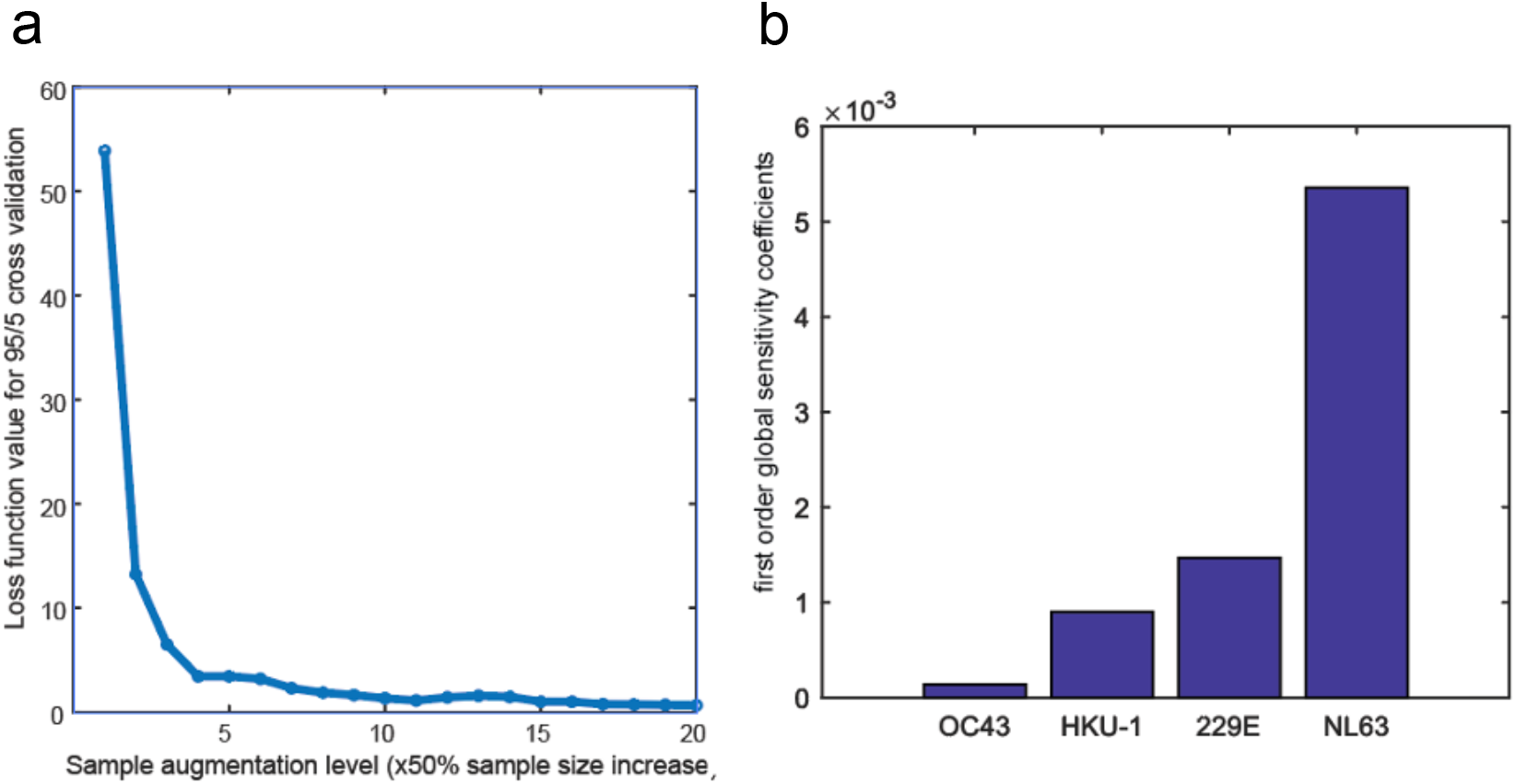
(a) The effect of sample size increase was determined using up-sampling with SMOTE algorithm on the loss function in the cross-validation study of our GPR model showing major increase in cross validation accuracy (reduced Loss function) with sample size increase. Loss function is calculated as: 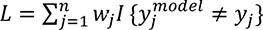 where *w_j_* is the weight for observation *j* normalized so that they sum to the corresponding prior class probability. (b) Global sensitivity analysis showing the contribution of the uncertainty of each input factor on the uncertainty of the output. (Calculations were performed using library FLAX running under Matlab (2021). Global Sensitivity Analysis Toolbox; MATLAB Central File Exchange. Retrieved September 25, 2021). Shown are first order global sensitivity coefficients calculated for 30,000 samples for the quasi-random Monte Carlo sampling calculated using Fourier amplitude sensitivity test (FAST) (Saltelli, A., and Bolado, R. (1998). An alternative way to compute Fourier amplitude sensitivity test (FAST). *Comput. Stat. Data Anal.* 26, 445–460. doi: 10.1016/S0167-9473(97)00043-1).

**Table S1.**
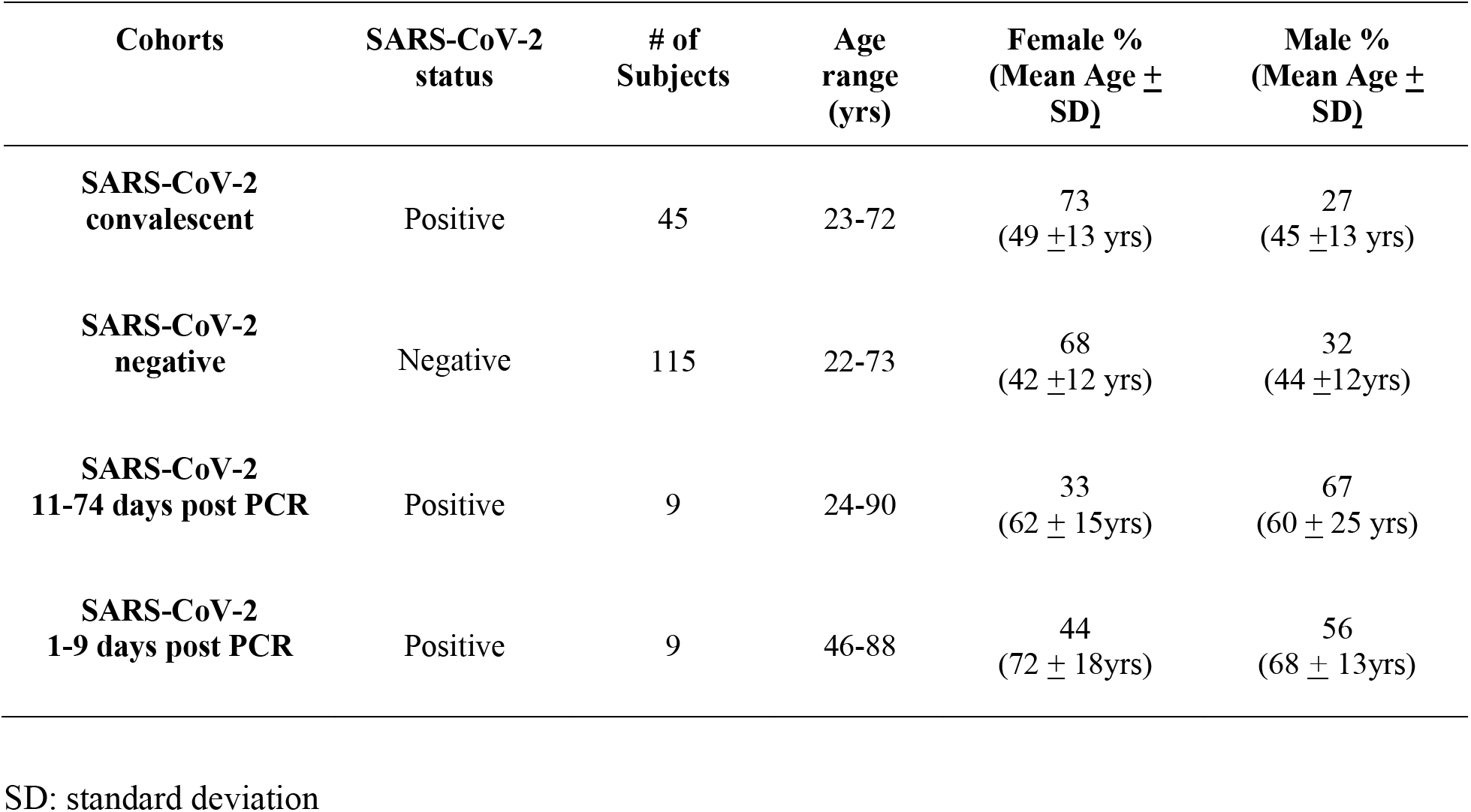
Demographic information of post-pandemic controls. Serum samples collected in various longitudinal studies of SARS-CoV-2 infected and non-infected individuals were used as reference groups. Demographic information was collected from the respective study or patient file and compiled.

| Cohorts | SARS-CoV-2 status | # of Subjects | Age range (yrs) | Female % (Mean Age ± SD) | Male % (Mean Age ± SD) |
| --- | --- | --- | --- | --- | --- |
| **SARS-CoV-2 convalescent** | Positive | 45 | 23-72 | 73<br>(49 ± 13 yrs) | 27<br>(45 ± 13 yrs) |
| **SARS-CoV-2 negative** | Negative | 115 | 22-73 | 68<br>(42 ± 12 yrs) | 32<br>(44 ± 12 yrs) |
| **SARS-CoV-2 11-74 days post PCR** | Positive | 9 | 24-90 | 33<br>(62 ± 15 yrs) | 67<br>(60 ± 25 yrs) |
| **SARS-CoV-2 1-9 days post PCR** | Positive | 9 | 46-88 | 44<br>(72 ± 18 yrs) | 56<br>(68 ± 13 yrs) |

SD: standard deviation

**Table S2.**
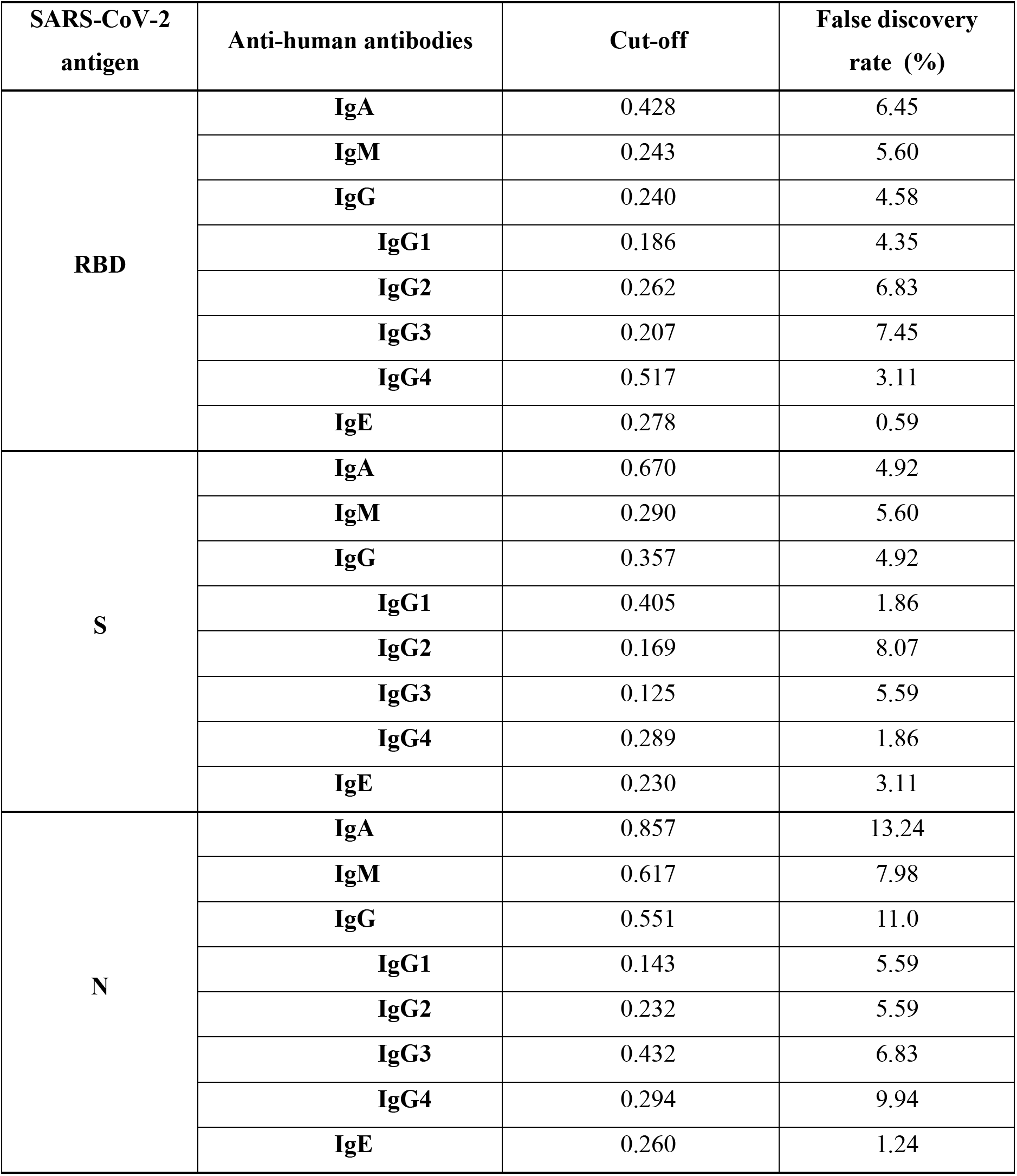
Determination of SARS-CoV-2 assay cut-off values of the four cohorts combined and false discovery rate (FDR) per antigen, isotype & subtypes. Cut-off values were calculated from two exclusion cycles at 2 standard deviations from the negative distributions. Any sample above that designated cut-off was included to determine the FDR

| **SARS-CoV-2 antigen** | **Anti-human antibodies** | **Cut-off** | **False discovery rate (%)** |
| --- | --- | --- | --- |
| **RBD** | **IgA** | 0.428 | 6.45 |
|  | **IgM** | 0.243 | 5.60 |
|  | **IgG** | 0.240 | 4.58 |
|  | **IgG1** | 0.186 | 4.35 |
|  | **IgG2** | 0.262 | 6.83 |
|  | **IgG3** | 0.207 | 7.45 |
|  | **IgG4** | 0.517 | 3.11 |
|  | **IgE** | 0.278 | 0.59 |
| **S** | **IgA** | 0.670 | 4.92 |
|  | **IgM** | 0.290 | 5.60 |
|  | **IgG** | 0.357 | 4.92 |
|  | **IgG1** | 0.405 | 1.86 |
|  | **IgG2** | 0.169 | 8.07 |
|  | **IgG3** | 0.125 | 5.59 |
|  | **IgG4** | 0.289 | 1.86 |
|  | **IgE** | 0.230 | 3.11 |
| **N** | **IgA** | 0.857 | 13.24 |
|  | **IgM** | 0.617 | 7.98 |
|  | **IgG** | 0.551 | 11.0 |
|  | **IgG1** | 0.143 | 5.59 |
|  | **IgG2** | 0.232 | 5.59 |
|  | **IgG3** | 0.432 | 6.83 |
|  | **IgG4** | 0.294 | 9.94 |
|  | **IgE** | 0.260 | 1.24 |

**Table S3.** Determination of SARS-CoV-2 assay cut-off values of pre-COVID-19 HCV and HIV patients and corresponding false discovery rate per antigen, isotype & subtypes. Cut-off values were calculated from two exclusion cycles at 2 standard deviations from the negative distributions. Any sample above that designated cut-off was included to determine the FDR.

| **SARS-CoV-2 antigen** | **Anti-human antibodies** | **Cut-off** | **False discovery rate (%)** |
| --- | --- | --- | --- |
| **RBD** | **IgA** | 0.428 | 6.78 |
|  | **IgM** | 0.243 | 5.08 |
|  | **IgG** | 0.240 | 8.47 |
|  | **IgG1** | 0.186 | 10.7 |
|  | **IgG2** | 0.262 | 0.00 |
|  | **IgG3** | 0.207 | 21.4 |
|  | **IgG4** | 0.517 | 0.00 |
|  | **IgE** | 0.278 | 0.00 |
| **S** | **IgA** | 0.670 | 13.6 |
|  | **IgM** | 0.290 | 6.78 |
|  | **IgG** | 0.357 | 6.78 |
|  | **IgG1** | 0.405 | 7.14 |
|  | **IgG2** | 0.169 | 7.14 |
|  | **IgG3** | 0.125 | 0.00 |
|  | **IgG4** | 0.289 | 0.00 |
|  | **IgE** | 0.230 | 7.14 |
| **N** | **IgA** | 0.857 | 15.3 |
|  | **IgM** | 0.617 | 6.78 |
|  | **IgG** | 0.551 | 15.3 |
|  | **IgG1** | 0.143 | 3.57 |
|  | **IgG2** | 0.232 | 3.57 |
|  | **IgG3** | 0.432 | 14.3 |
|  | **IgG4** | 0.294 | 17.9 |
|  | **IgE** | 0.260 | 3.57 |

**Table S4.** Determination of SARS-CoV-2 assay cut-off values of pre-COVID-19 > 70 yrs of age and corresponding false discovery rate per antigen, isotype & subtypes. Cut-off values were calculated from two exclusion cycles at 2 standard deviations from the negative distributions. Any sample above that designated cut-off was included to determine the FDR.

| **SARS-CoV-2 antigen** | **Anti-human antibodies** | **Cut-off** | **False discovery rate (%)** |
| --- | --- | --- | --- |
| **RBD** | **IgA** | 0.428 | 6.25 |
|  | **IgM** | 0.243 | 3.13 |
|  | **IgG** | 0.240 | 1.56 |
|  | **IgG1** | 0.186 | 0.00 |
|  | **IgG2** | 0.262 | 8.33 |
|  | **IgG3** | 0.207 | 0.00 |
|  | **IgG4** | 0.517 | 0.00 |
|  | **IgE** | 0.278 | 8.33 |
| **S** | **IgA** | 0.670 | 1.56 |
|  | **IgM** | 0.290 | 0.00 |
|  | **IgG** | 0.357 | 1.56 |
|  | **IgG1** | 0.405 | 0.00 |
|  | **IgG2** | 0.169 | 16.7 |
|  | **IgG3** | 0.125 | 0.00 |
|  | **IgG4** | 0.289 | 0.00 |
|  | **IgE** | 0.230 | 0.00 |
| **N** | **IgA** | 0.857 | 21.9 |
|  | **IgM** | 0.617 | 6.25 |
|  | **IgG** | 0.551 | 14.1 |
|  | **IgG1** | 0.143 | 8.33 |
|  | **IgG2** | 0.232 | 0.00 |
|  | **IgG3** | 0.432 | 8.33 |
|  | **IgG4** | 0.294 | 8.33 |
|  | **IgE** | 0.260 | 0.00 |

**Table S5.**
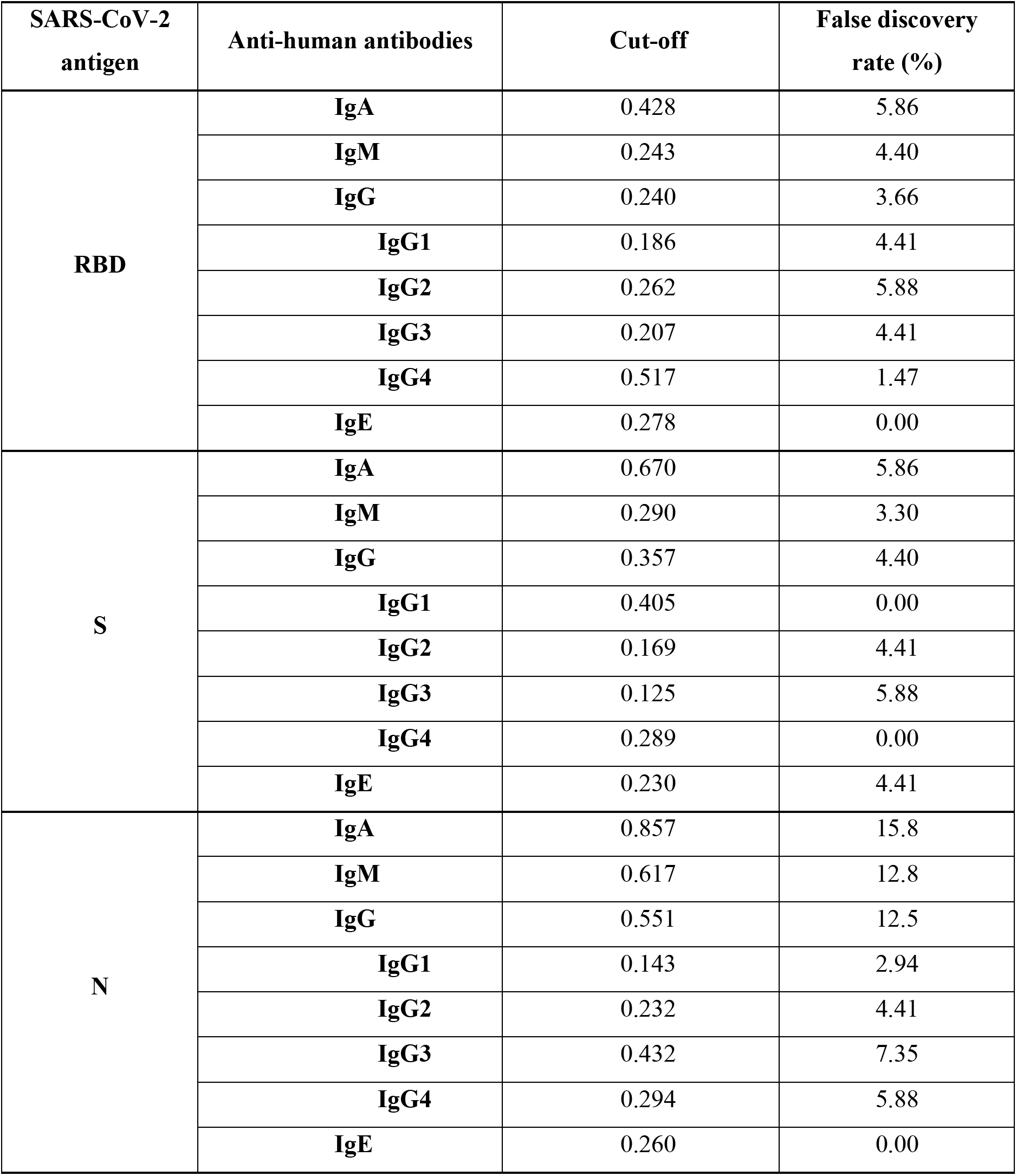
Determination of SARS-CoV-2 assay cut-off values of pre-COVID-19 21 - 70 yrs of age and corresponding false discovery rate per antigen, isotype & subtypes. Cut-off values were calculated from two exclusion cycles at 2 standard deviations from the negative distributions. Any sample above that designated cut-off was included to determine the FDR.

**Table S6.** Determination of SARS-CoV-2 assay cut-off values of pre-COVID 19 < 21 yrs of age and corresponding false discovery rate per antigen, isotype & subtypes. Cut-off values were calculated from two exclusion cycles at 2 standard deviations from the negative distributions. Any sample above that designated cut-off was included to determine the FDR.

| **SARS-CoV-2 antigen** | **Anti-human antibodies** | **Cut-off** | **False discovery rate (%)** |
| --- | --- | --- | --- |
| **RBD** | **IgA** | 0.428 | 7.25 |
|  | **IgM** | 0.243 | 8.29 |
|  | **IgG** | 0.240 | 5.70 |
|  | **IgG1** | 0.186 | 1.89 |
|  | **IgG2** | 0.262 | 11.3 |
|  | **IgG3** | 0.207 | 5.66 |
|  | **IgG4** | 0.517 | 7.55 |
|  | **IgE** | 0.278 | 0.00 |
| **S** | **IgA** | 0.670 | 2.07 |
|  | **IgM** | 0.290 | 10.4 |
|  | **IgG** | 0.357 | 6.22 |
|  | **IgG1** | 0.405 | 1.89 |
|  | **IgG2** | 0.169 | 11.3 |
|  | **IgG3** | 0.125 | 9.43 |
|  | **IgG4** | 0.289 | 3.77 |
|  | **IgE** | 0.230 | 0.00 |
| **N** | **IgA** | 0.857 | 6.22 |
|  | **IgM** | 0.617 | 2.07 |
|  | **IgG** | 0.551 | 6.74 |
|  | **IgG1** | 0.143 | 9.43 |
|  | **IgG2** | 0.232 | 9.43 |
|  | **IgG3** | 0.432 | 1.89 |
|  | **IgG4** | 0.294 | 11.3 |
|  | **IgE** | 0.260 | 1.89 |

**Table S7.** Multi-antigen FDR calculation. False discovery rate of all four cohorts combined using a single antigen, combinations of two antigens and three antigens were compared by each isotype and subtype.

| Antibody<br>Isotype | Single antigen FDR<br>(%) |  |  | Two antigen FDR<br>(%) |  |  | Three antigen FDR<br>(%) |
| --- | --- | --- | --- | --- | --- | --- | --- |
|  | RBD | Spike | N | RBD<br>+ S | RBD<br>+ N | S<br>+ N | RBD + S+N |
| IgA | 6.45 | 4.92 | 13.2 | 1.19 | 1.87 | 1.70 | 0.68 |
| IgM | 5.60 | 5.60 | 7.98 | 2.21 | 1.19 | 0.68 | 0.51 |
| IgG | 4.58 | 4.92 | 11.0 | 1.53 | 1.53 | 0.85 | 0.51 |
| IgG1 | 4.35 | 1.86 | 5.59 | 0.62 | 0.62 | 0.00 | 0.00 |
| IgG2 | 6.83 | 8.07 | 5.59 | 0.62 | 0.62 | 0.00 | 0.00 |
| IgG3 | 7.45 | 5.59 | 6.83 | 0.62 | 1.86 | 0.62 | 0.00 |
| IgG4 | 3.11 | 1.86 | 9.94 | 0.62 | 0.62 | 0.62 | 0.62 |
| IgE | 0.62 | 3.11 | 1.24 | 0.00 | 0.00 | 0.00 | 0.00 |

**Table S8.** Demographics of fuzzy mean clusters of patients segregated by serum antibody cross-reactivity. Cross-reactivity against 3 different SARS-CoV-2 antigens (N, RBD, Spike) for IgA, IgM and IgG were included with capacity of these antibodies to inhibit Spike-ACE2 binding.

| **Cluster #**<br>**(n)\*** | **Age**<br>**Mean ± SD**<br>**(Range)** | **Sex**<br>**(% Female)** |
| --- | --- | --- |
| 1 (79) | 41±26 (1-90 yrs) | 58 |
| 2 (21) | 32±23 (3-69 yrs) | 42 |
| 3 (16) | 45±24 (4-68 yrs) | 38 |
| 4 (36) | 45±26 (1-79 yrs) | 44 |
| 5 (86) | 45±26 (1-90 yrs) | 47 |
| 6 (1) | 21 yrs | 0 |
| 7 (53) | 41±28 (2-80 yrs) | 42 |
| 8 (89) | 35±28 (1-90 yrs) | 47 |
| 9 (65) | 35±28 (2-96 yrs) | 57 |
| 10 (60) | 36±28 (1-97 yrs) | 50 |

Total n = 507\*.

## References

1. Anderson EM, Goodwin EC, Verma A, Arevalo CP, Bolton MJ, Weirick ME, et al. Seasonal human coronavirus antibodies are boosted upon SARS-CoV-2 infection but not associated with protection. Cell. 2021;184(7):1858–64.e10.

2. Gaunt ER, Hardie A, Claas EC, Simmonds P, Templeton KE. Epidemiology and clinical presentations of the four human coronaviruses 229E, HKU1, NL63, and OC43 detected over 3 years using a novel multiplex real-time PCR method. J Clin Microbiol. 2010;48(8):2940–7.

3. Edridge AWD, Kaczorowska J, Hoste ACR, Bakker M, Klein M, Loens K, et al. Seasonal coronavirus protective immunity is short-lasting. Nat Med. 2020;26(11):1691–3.

4. Masse S, Capai L, Villechenaud N, Blanchon T, Charrel R, Falchi A. Epidemiology and Clinical Symptoms Related to Seasonal Coronavirus Identified in Patients with Acute Respiratory Infections Consulting in Primary Care over Six Influenza Seasons (2014-2020) in France. Viruses. 2020;12(6).

5. Gorse GJ, Patel GB, Vitale JN, O’Connor TZ. Prevalence of Antibodies to Four Human Coronaviruses Is Lower in Nasal Secretions than in Serum. Clinical and Vaccine Immunology. 2010;17(12):1875–80.

6. Song G, He WT, Callaghan S, Anzanello F, Huang D, Ricketts J, et al. Cross-reactive serum and memory B-cell responses to spike protein in SARS-CoV-2 and endemic coronavirus infection. Nat Commun. 2021;12(1):2938.

7. Majdoubi A, Michalski C, O’Connell SE, Dada S, Narpala S, Gelinas J, et al. A majority of uninfected adults show preexisting antibody reactivity against SARS-CoV-2. JCI Insight. 2021;6(8).

8. V’kovski P, Kratzel A, Steiner S, Stalder H, Thiel V. Coronavirus biology and replication: implications for SARS-CoV-2. Nature Reviews Microbiology. 2021;19(3):155–70.

9. Hicks J, Klumpp-Thomas C, Kalish H, Shunmugavel A, Mehalko J, Denson JP, et al. Serologic Cross-Reactivity of SARS-CoV-2 with Endemic and Seasonal Betacoronaviruses. J Clin Immunol. 2021;41(5):906–13.

10. Grzelak L, Temmam S, Planchais C, Demeret C, Tondeur L, Huon C, et al. A comparison of four serological assays for detecting anti-SARS-CoV-2 antibodies in human serum samples from different populations. Sci Transl Med. 2020;12(559).

11. Okba NMA, Muller MA, Li W, Wang C, GeurtsvanKessel CH, Corman VM, et al. Severe Acute Respiratory Syndrome Coronavirus 2-Specific Antibody Responses in Coronavirus Disease Patients. Emerg Infect Dis. 2020;26(7):1478–88.

12. Lv H, Wu NC, Tsang OT, Yuan M, Perera R, Leung WS, et al. Cross-reactive Antibody Response between SARS-CoV-2 and SARS-CoV Infections. Cell Rep. 2020;31(9):107725.

13. Ladner JT, Henson SN, Boyle AS, Engelbrektson AL, Fink ZW, Rahee F, et al. Epitope-resolved profiling of the SARS-CoV-2 antibody response identifies cross-reactivity with endemic human coronaviruses. Cell Rep Med. 2021;2(1):100189.

14. Khan T, Rahman M, Ali FA, Huang SSY, Ata M, Zhang Q, et al. Distinct antibody repertoires against endemic human coronaviruses in children and adults. JCI Insight. 2021;6(4).

15. Trivedi SU, Miao C, Sanchez JE, Caidi H, Tamin A, Haynes L, et al. Development and Evaluation of a Multiplexed Immunoassay for Simultaneous Detection of Serum IgG Antibodies to Six Human Coronaviruses. Scientific Reports. 2019;9(1):1390.

16. Galipeau Y, Greig M, Liu G, Driedger M, Langlois M-A. Humoral Responses and Serological Assays in SARS-CoV-2 Infections. Frontiers in Immunology. 2020;11(3382).

17. Piccoli L, Park YJ, Tortorici MA, Czudnochowski N, Walls AC, Beltramello M, et al. Mapping Neutralizing and Immunodominant Sites on the SARS-CoV-2 Spike Receptor-Binding Domain by Structure-Guided High-Resolution Serology. Cell. 2020;183(4):1024–42.e21.

18. McCallum M, De Marco A, Lempp FA, Tortorici MA, Pinto D, Walls AC, et al. N-terminal domain antigenic mapping reveals a site of vulnerability for SARS-CoV-2. Cell. 2021;184(9):2332–47.e16.

19. Andreano E, Nicastri E, Paciello I, Pileri P, Manganaro N, Piccini G, et al. Extremely potent human monoclonal antibodies from COVID-19 convalescent patients. Cell. 2021;184(7):1821–35.e16.

20. Graham C, Seow J, Huettner I, Khan H, Kouphou N, Acors S, et al. Neutralization potency of monoclonal antibodies recognizing dominant and subdominant epitopes on SARS-CoV-2 Spike is impacted by the B.1.1.7 variant. Immunity. 2021;54(6):1276–89.e6.

21. McMahan K, Yu J, Mercado NB, Loos C, Tostanoski LH, Chandrashekar A, et al. Correlates of protection against SARS-CoV-2 in rhesus macaques. Nature. 2021;590(7847):630–4.

22. Anand SP, Prevost J, Nayrac M, Beaudoin-Bussieres G, Benlarbi M, Gasser R, et al. Longitudinal analysis of humoral immunity against SARS-CoV-2 Spike in convalescent individuals up to 8 months post-symptom onset. Cell Rep Med. 2021;2(6):100290.

23. McAndrews KM, Dowlatshahi DP, Dai J, Becker LM, Hensel J, Snowden LM, et al. Heterogeneous antibodies against SARS-CoV-2 spike receptor binding domain and nucleocapsid with implications for COVID-19 immunity. JCI Insight. 2020;5(18).

24. Grifoni A, Weiskopf D, Ramirez SI, Mateus J, Dan JM, Moderbacher CR, et al. Targets of T Cell Responses to SARS-CoV-2 Coronavirus in Humans with COVID-19 Disease and Unexposed Individuals. Cell. 2020;181(7):1489–501.e15.

25. Braun J, Loyal L, Frentsch M, Wendisch D, Georg P, Kurth F, et al. SARS-CoV-2-reactive T cells in healthy donors and patients with COVID-19. Nature. 2020;587(7833):270–4.

26. Le Bert N, Tan AT, Kunasegaran K, Tham CYL, Hafezi M, Chia A, et al. SARS-CoV-2-specific T cell immunity in cases of COVID-19 and SARS, and uninfected controls. Nature. 2020;584(7821):457–62.

27. Anderson DE, Tan CW, Chia WN, Young BE, Linster M, Low JH, et al. Lack of cross-neutralization by SARS patient sera towards SARS-CoV-2. Emerging Microbes & Infections. 2020;9(1):900–2.

28. Prévost J, Gasser R, Beaudoin-Bussières G, Richard J, Duerr R, Laumaea A, et al. Cross-Sectional Evaluation of Humoral Responses against SARS-CoV-2 Spike. Cell Reports Medicine. 2020;1(7).

29. Sagar M, Reifler K, Rossi M, Miller NS, Sinha P, White LF, et al. Recent endemic coronavirus infection is associated with less-severe COVID-19. The Journal of Clinical Investigation. 2021;131(1).

30. Huang AT, Garcia-Carreras B, Hitchings MDT, Yang B, Katzelnick LC, Rattigan SM, et al. A systematic review of antibody mediated immunity to coronaviruses: kinetics, correlates of protection, and association with severity. Nature Communications. 2020;11(1):4704.

31. Poston D, Weisblum Y, Wise H, Templeton K, Jenks S, Hatziioannou T, et al. Absence of Severe Acute Respiratory Syndrome Coronavirus 2 Neutralizing Activity in Prepandemic Sera From Individuals With Recent Seasonal Coronavirus Infection. Clinical Infectious Diseases. 2020.

32. Morgenlander WR, Henson SN, Monaco DR, Chen A, Littlefield K, Bloch EM, et al. Antibody responses to endemic coronaviruses modulate COVID-19 convalescent plasma functionality. J Clin Invest. 2021;131(7).

33. Schulien I, Kemming J, Oberhardt V, Wild K, Seidel LM, Killmer S, et al. Characterization of pre-existing and induced SARS-CoV-2-specific CD8(+) T cells. Nat Med. 2021;27(1):78–85.

34. Mateus J, Grifoni A, Tarke A, Sidney J, Ramirez SI, Dan JM, et al. Selective and cross-reactive SARS-CoV-2 T cell epitopes in unexposed humans. Science. 2020;370(6512):89–94.

35. Henss L, Scholz T, von Rhein C, Wieters I, Borgans F, Eberhardt FJ, et al. Analysis of Humoral Immune Responses in Patients With Severe Acute Respiratory Syndrome Coronavirus 2 Infection. J Infect Dis. 2021;223(1):56–61.

36. Székely GJ, Rizzo ML, Bakirov NK. Measuring and testing dependence by correlation of distances. The Annals of Statistics. 2007;35(6):2769–94, 26.

37. Robnik-Šikonja M, Kononenko I. Theoretical and Empirical Analysis of ReliefF and RReliefF. Machine Learning. 2003;53(1):23–69.

38. Poulain A, Perret S, Malenfant F, Mullick A, Massie B, Durocher Y. Rapid protein production from stable CHO cell pools using plasmid vector and the cumate gene-switch. J Biotechnol. 2017;255:16–27.

39. Wevers BA, van der Hoek L. Recently discovered human coronaviruses. Clin Lab Med. 2009;29(4):715–24.

40. Stadlbauer D, Amanat F, Chromikova V, Jiang K, Strohmeier S, Arunkumar GA, et al. SARS-CoV-2 Seroconversion in Humans: A Detailed Protocol for a Serological Assay, Antigen Production, and Test Setup. Current Protocols in Microbiology. 2020;57(1):e100.

41. Isho B, Abe KT, Zuo M, Jamal AJ, Rathod B, Wang JH, et al. Persistence of serum and saliva antibody responses to SARS-CoV-2 spike antigens in COVID-19 patients. Science Immunology. 2020;5(52):eabe5511.

42. Leuridan E, Hens N, Hutse V, Aerts M, Van Damme P. Kinetics of maternal antibodies against rubella and varicella in infants. Vaccine. 2011;29(11):2222–6.

43. Leuridan E, Van Damme P. Passive transmission and persistence of naturally acquired or vaccine-induced maternal antibodies against measles in newborns. Vaccine. 2007;25(34):6296–304.

44. Niewiesk S. Maternal antibodies: clinical significance, mechanism of interference with immune responses, and possible vaccination strategies. Front Immunol. 2014;5:446.

45. Dijkman R, Jebbink MF, Gaunt E, Rossen JW, Templeton KE, Kuijpers TW, et al. The dominance of human coronavirus OC43 and NL63 infections in infants. J Clin Virol. 2012;53(2):135–9.

46. Severance EG, Bossis I, Dickerson FB, Stallings CR, Origoni AE, Sullens A, et al. Development of a Nucleocapsid-Based Human Coronavirus Immunoassay and Estimates of Individuals Exposed to Coronavirus in a U.S. Metropolitan Population. Clinical and Vaccine Immunology. 2008;15(12):1805–10.

47. Dijkman R, Jebbink MF, El Idrissi NB, Pyrc K, Müller MA, Kuijpers TW, et al. Human Coronavirus NL63 and 229E Seroconversion in Children. Journal of Clinical Microbiology. 2008;46(7):2368–73.

48. van Tol S, Mögling R, Li W, Godeke G-J, Swart A, Bergmans B, et al. Accurate serology for SARS-CoV-2 and common human coronaviruses using a multiplex approach. Emerging Microbes & Infections. 2020;9(1):1965–73.

49. Ahmed SF, Quadeer AA, McKay MR. Preliminary Identification of Potential Vaccine Targets for the COVID-19 Coronavirus (SARS-CoV-2) Based on SARS-CoV Immunological Studies. Viruses. 2020;12(3).

50. Algaissi A, Alfaleh MA, Hala S, Abujamel TS, Alamri SS, Almahboub SA, et al. SARS-CoV-2 S1 and N-based serological assays reveal rapid seroconversion and induction of specific antibody response in COVID-19 patients. Scientific Reports. 2020;10(1):16561.

51. Grzelak L, Temmam S, Planchais C, Demeret C, Tondeur L, Huon C, et al. A comparison of four serological assays for detecting anti–SARS-CoV-2 antibodies in human serum samples from different populations. Science Translational Medicine. 2020;12(559):eabc3103.

52. Amarasekera M. Immunoglobulin E in health and disease. Asia Pac Allergy. 2011;1(1):12–5.

53. Goh YS, Chavatte J-M, Lim Jieling A, Lee B, Hor PX, Amrun SN, et al. Sensitive detection of total anti-Spike antibodies and isotype switching in asymptomatic and symptomatic individuals with COVID-19. Cell Reports Medicine.

54. Abe KT, Li Z, Samson R, Samavarchi-Tehrani P, Valcourt EJ, Wood H, et al. A simple protein-based surrogate neutralization assay for SARS-CoV-2. JCI Insight. 2020;5(19).

55. Wang Y, Zhang L, Sang L, Ye F, Ruan S, Zhong B, et al. Kinetics of viral load and antibody response in relation to COVID-19 severity. The Journal of Clinical Investigation. 2020;130(10).

56. Liu L, Wang P, Nair MS, Yu J, Rapp M, Wang Q, et al. Potent neutralizing antibodies against multiple epitopes on SARS-CoV-2 spike. Nature. 2020;584(7821):450–6.

57. Brouwer PJM, Caniels TG, van der Straten K, Snitselaar JL, Aldon Y, Bangaru S, et al. Potent neutralizing antibodies from COVID-19 patients define multiple targets of vulnerability. Science. 2020;369(6504):643–50.

58. Zost SJ, Gilchuk P, Chen RE, Case JB, Reidy JX, Trivette A, et al. Rapid isolation and profiling of a diverse panel of human monoclonal antibodies targeting the SARS-CoV-2 spike protein. Nature Medicine. 2020;26(9):1422–7.

59. Selva KJ, van de Sandt CE, Lemke MM, Lee CY, Shoffner SK, Chua BY, et al. Systems serology detects functionally distinct coronavirus antibody features in children and elderly. Nature Communications. 2021;12(1):2037.

60. Ghosh S, Dellibovi-Ragheb TA, Kerviel A, Pak E, Qiu Q, Fisher M, et al. beta-Coronaviruses Use Lysosomes for Egress Instead of the Biosynthetic Secretory Pathway. Cell. 2020;183(6):1520–35 e14.

61. V’Kovski P, Kratzel A, Steiner S, Stalder H, Thiel V. Coronavirus biology and replication: implications for SARS-CoV-2. Nat Rev Microbiol. 2021;19(3):155–70.

62. Dugas M, Grote-Westrick T, Vollenberg R, Lorentzen E, Brix T, Schmidt H, et al. Less severe course of COVID-19 is associated with elevated levels of antibodies against seasonal human coronaviruses OC43 and HKU1 (HCoV OC43, HCoV HKU1). Int J Infect Dis. 2021;105:304–6.

63. Shrock E, Fujimura E, Kula T, Timms RT, Lee I-H, Leng Y, et al. Viral epitope profiling of COVID-19 patients reveals cross-reactivity and correlates of severity. Science. 2020;370(6520):eabd4250.

